# On real-time calibrated prediction for complex model-based decision support in pandemics: Part 1

**DOI:** 10.1101/2025.05.16.25327688

**Authors:** Daniel B. Williamson, Trevelyan J. McKinley, Xiaoyu Xiong, James M. Salter, Robert Challen, Leon Danon, Benjamin D. Youngman, Doug McNeall

**Affiliations:** Department of Mathematics and Statistics, University of Exeter, Exeter, UK; University of Exeter Medical School, University of Exeter, Exeter, UK; Department of Engineering Mathematics, University of Bristol, Bristol, UK; UK Met Office, Exeter, UK

**Keywords:** Model Discrepancy, Calibration, Data Assimilation, COVID-19, Deep Gaussian Process

## Abstract

Infectious disease models are used to predict the spread and impact of outbreaks of a disease. Like other complex models, they have parameters that need to be calibrated, and structural discrepancies from the reality that they simulate that should be accounted for in calibration and prediction. Whilst Uncertainty Quantification (UQ) techniques have been applied to infectious disease models before, they were not routinely used to inform policymakers in the UK during the COVID-19 pandemic. In this paper, we will argue that during a fast moving pandemic, models and policy are changing on timescales that make traditional UQ methods impractical, if not impossible to implement. We present an alternative formulation to the calibration problem that embeds model discrepancy within the structure of the model, and appropriately assimilates data within the simulation. We then show how UQ can be used to calibrate the model in real-time to produce disease trajectories accounting for parameter uncertainty and model discrepancy. We apply these ideas to an age-structured COVID-19 model for England and demonstrate the types of information it could have produced to feed into policy support prior to the lockdown of March 2020.

## 1 Introduction

Infectious disease (ID) models are widely used by epidemiologists and public health officials to predict the spread and impact of outbreaks of a disease, notably during the recent COVID-19 pandemic [e.g. Brooks-Pollock et al., 2021, Panovska-Griffiths et al., 2022, Kretzschmar et al., 2022, Ackland et al., 2022]. By mathematically representing transmission dynamics they can provide insights into how interventions (like vaccinations, social distancing, or quarantines) might affect an outbreak, as well as providing quantitative estimates for key epidemiological parameters and predictions/forecasts of the epidemic trajectories, as long as they can be robustly fitted to incomplete data. ID models range from simple deterministic models like the classic SIR (Susceptible-Infectious-Recovered) model (which treats the number of individuals in each epidemiological state as a set of continuous quantities and then models the flow of individuals between states through a set of coupled differential equations), right through to stochastic agent-based models where epidemiological dynamics are modelled through probabilistic interactions between individuals within a population. These models can be extended to add further complexities including age and spatial structure, and even multiple interacting diseases [e.g Danon et al., 2009, Aylett-Bullock et al., 2021].

Methods under the collective umbrella of ‘Uncertainty Quantification’ (UQ), have long been used to quantify the uncertainties present when using computationally expensive simulation models to learn about the real world [Williamson et al., 2017, Bingham et al., 2024, Owen et al., 2017]. Unfortunately, the name ‘Uncertainty Quantification’ itself is not particularly useful, as it can often be misunderstood. For example, to say that uncertainty quantification is not routinely used in ID modelling, is both correct, from the lens of those within the UQ sub-discipline, and incorrect and perhaps derogatory from a literal reading of the words ‘uncertainty quantification’. ID modellers trying to calibrate their models are routinely concerned with quantifying uncertainties, particularly the uncertainties present in the data they have for model calibration and how that is propagated to uncertainty in forecasts/projections of an epidemic [e.g. Read et al., 2021, Challen et al., 2021, Overton et al., 2020]. However, in this paper we use the term UQ to refer to a specific subdiscipline within statistics and machine learning, where its principal contributions include: emulation / surrogate modelling (that is, providing fast statistical models of the slower simulation models for use within algorithms) [Rasmussen and Williams, 2006, Gramacy, 2020, Ming et al., 2023]; calibration (using emulators and quantified model discrepancy to estimate model parameters using data from the system that is being simulated) [Kennedy and O’Hagan, 2001, Higdon et al., 2008, Vernon et al., 2010, Andrianakis et al., 2015, Hourdin et al., 2023]; as well as optimisation and sensitivity analysis [Oakley and O’Hagan, 2004, Gramacy and Lee, 2011]. UQ approaches, particularly those utilising emulators, have been shown to be very useful for inference and prediction for any model (ID models included), but particularly for computationally intensive models. Furthermore, UQ-practitioners argue that incorporating model discrepancy is essential, particularly for calibration [Brynjarsdóttir and O’Hagan, 2014, Dunne et al., 2022, Swallow et al., 2022].

During the recent COVID-19 pandemic in the UK, many ID models were used to forecast the evolution of the outbreak, its impact on hospital and ventilator capacity, numbers of deaths and efficacy of potential policy interventions, including lockdown and vaccination rollout [e.g. Ferguson et al., 2020, Keeling et al., 2021, Birrell et al., 2021, Danon et al., 2021, Hill et al., 2021, Moore et al., 2021, Manley et al., 2021, Silk et al., 2022, Bowman, 2022, Kretzschmar et al., 2022]. Several respected modelling groups were invited to participate in the Scientific Pandemic Influenza Group on Modelling, Operational subgroup (SPI-M-O) of the Scientific Advisory Group for Emergencies (SAGE), the body responsible for giving expert advice to government. These groups used their modelling frameworks and sensitive data streams to provide forecasts (with uncertainty) during the pandemic and for different policy scenarios [see e.g. Brooks-Pollock et al., 2021]. Despite the use of highly sophisticated statistical techniques by many of these groups, UQ methods, deemed both useful and critical (for calibration) by our field, were not routinely used.

UQ has been applied to ID models in a number of previous studies [e.g. Andrianakis et al., 2015, 2017, McCreesh et al., 2017a,b, McKinley et al., 2018, Fadikar et al., 2018], and during the pandemic UQ tutorials were developed for epidemic models [Dunne et al., 2022] and case studies were published applying UQ to COVID-19 models [Swallow et al., 2022, Vernon et al., 2022, Edeling et al., 2021]. However, a key reason that UQ methods were not deployed in support of UK policymakers during the pandemic was that, in their off-the-shelf form, they were infeasible to implement within decision-relevant timescales. During the UQ4Covid project that funded this work, the author team, comprising UQ practitioners and ID modellers who were participating in SPI-M-O, attempted to deliver real-time UQ to feed into the policy support chain. ‘Real-time’ here typically meant a single day (with the data) and a week to prepare, during which the model itself would be adapted or changed to answer questions posed by policymakers. Whilst emulating any version of the model was always possible [see, e.g. Salter et al., 2025], it was not generally possible to do so effectively for calibration in a short time-scale, because the models were spatio-temporal and age-structured, and because the important questions (and hence ‘features’ of the model that were important) were always changing. These issues compounded for model discrepancy modelling and estimation, with additional challenges posed for some policy questions due to uncertainty in the initial state of the disease or any of its variants.

In this paper and its accompanying second part [McKinley et al., 2025], we present a framework to bring UQ methods into real-time modelling and policy support for epidemics. Our approach is to embed model discrepancy in all model states, for all locations and all age-classes, and to use particle filtering to approximate the likelihood for any data on the model states. With the discrepancy embedded, the rest of the UQ (emulation and calibration) is done on the log-likelihood, meaning that the UQ problem is 1D and can be automated. We have divided our presentation into two papers: Part 1 (this manuscript) describes the framework and develops the necessary innovations in UQ, with examples and an application to COVID-19 data for an age-structured but non-spatial model. Challenges posed by spatial models, particularly for the particle filtering with embedded discrepancy, are addressed in Part 2 [McKinley et al., 2025] in the context of a high-resolution application to modelling COVID-19 spread in England.

Part 1 is structured as follows: Section 2 describes traditional UQ methods within the context of stochastic ID modelling and outlines the challenges that applying these methods in real-time pandemic modelling pose. Section 3 presents data assimilation with embedded model discrepancy, including detailed modelling for the dependence structure of embedded discrepancy models in ID, and describes the calibration process using the resulting log-likelihood estimates. In Section 4 we present our age-structured model for COVID-19 in the lead up to lockdown in March 2020, and conduct some perfect model experiments to illustrate the methodology. Section 5 applies our framework to observations from the pandemic and performs scenario analysis for policymakers for the lockdown decision, to illustrate the types of analysis that can be turned around quickly with our framework. Section 6 contains discussion.

## 2 Traditional UQ vs. real-time calibration

We consider developing real-time uncertainty quantification for stochastic compartmental models of infectious disease transmission. As discussed earlier, this higher-level structure covers a very broad class of models, including agent-based models, though our COVID-19 application will use the stochastic model shown in Figure 1. Individuals begin in a susceptible (*S*) state and transition to an exposed (*E*) state according to model-dependent rules for transmission of the disease due to contact with infectious individuals (i.e. those in either *A, I*_1_ or *I*_2_). After being exposed, some individuals will transition down a pathway for asymptomatic infection (*A*), after which they will eventually recover (*R*_*A*_). Other individuals will transition down a symptomatic pathway, which includes a pre-symptomatic state (*P*) followed by a symptomatic infectious state (*I*_1_). From there, some individuals will transition to a recovering yet still infectious state (*I*_2_) before recovering completely (*R*_*I*_); some individuals will die (*D*_*I*_) and some individuals will transition to hospital (*H*), from which they will either eventually recover (*R*_*H*_) or die (*D*_*H*_). Due to the time-scales over which we are running the model we will assume that once recovered, individuals are immune to reinfection. More complex models could be built, where immunity wanes and eventually recovered individuals transition back to the susceptible class, but here we use the model described above to fix ideas.

**Figure 1:**
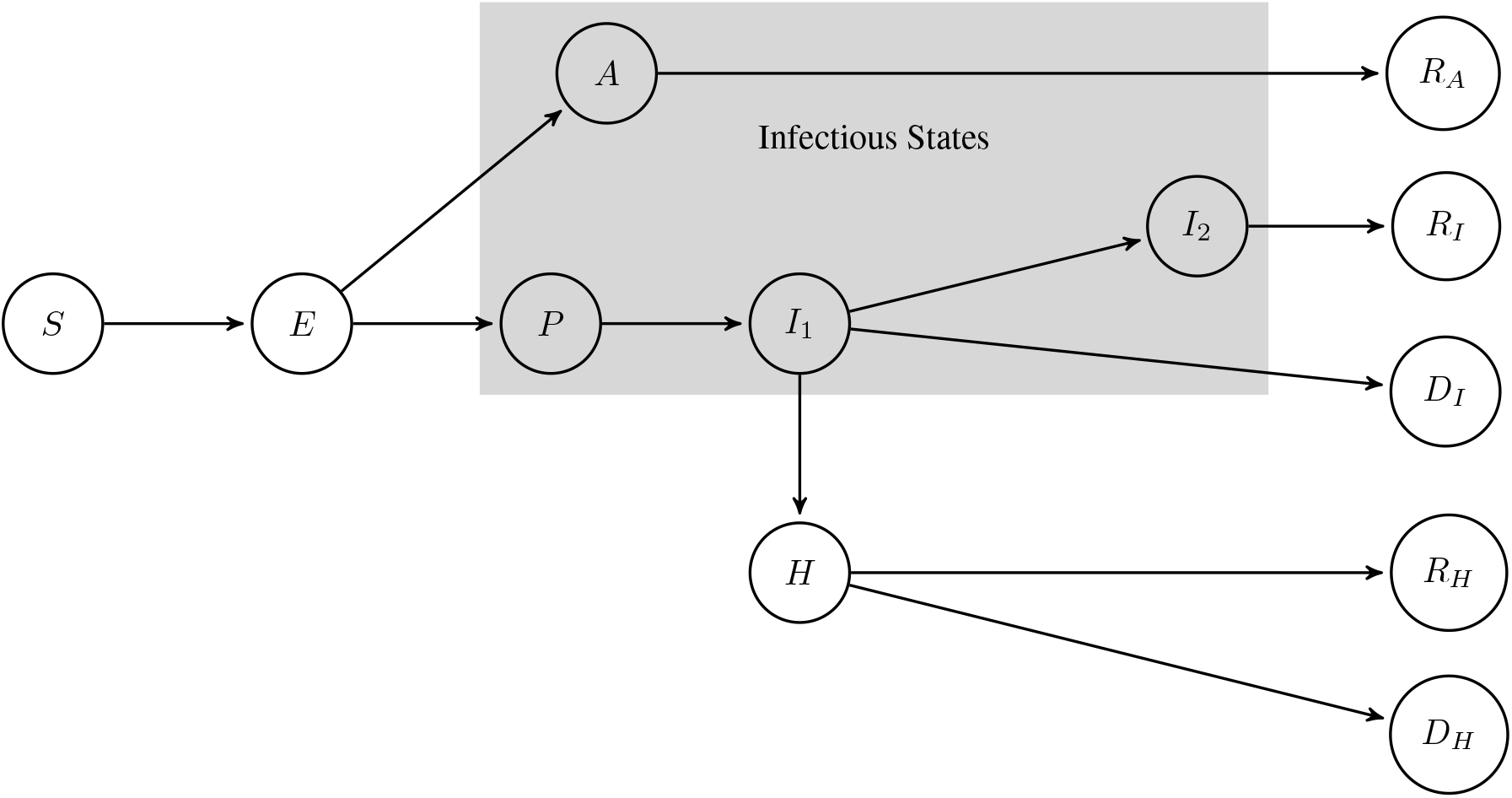
Schematic of the COVID-19 model considered in this paper and an example of the SEIR compartment structure of the class of infectious disease models we consider throughout.

Such a structure can also exist for different age-groups within a population (with different probabilities of moving down different pathways for each age group) and can be repeated for a large number of spatial locations simultaneously. The model itself will determine (usually probabilistic) rules for transition between states across different locations and these rules will themselves be governed by a number of parameters that must be input to the model.

### 2.1 Model discrepancy

To align with the infectious disease literature, we denote the state vector of an infectious disease model, ***X***, and make explicit that the state depends on a number of model parameters, which we denote ***θ***. In the example shown in Figure 1, ***X*** is the vector of counts (*S, E, A, P, I*_1_, *I*_2_, *H, R*_*A*_, *R*_*I*_, *D*_*I*_, *R*_*H*_, *D*_*H*_)^*T*^ for each age-class and each time point. We will use subscripts to denote the model state at time *t*, age-class *a* and state *c*, so that, for example, if *c* = *H* then the number in hospital in the simulation in age-class *a* at time *t* is *X*_*ta*,*H*_ . The parameter vector, ***θ***, will typically include disease parameters we want to infer, such as the basic reproduction number *R*_0_, and other model-specific parameters (see Section 5). In this paper we assume we are running the model in a single population, but in Part 2 [McKinley et al., 2025] we discuss extensions to deal with interacting spatial meta-populations.

A critical component of UQ approaches is to acknowledge *model discrepancy*, the idea that the state in the real world will not be equivalent to the model state, even if the parameters were known. UQ approaches make reality, data (reality observed with error) and the model distinct. Throughout we will denote the true state of reality via ***Y***, and our observations via ***Z***, using subscripts as discussed above to delinate states, age-classes and time points, noting that we will not generally observe the whole true state vector but only part of it (e.g. deaths aggregated over some region). Rather than write down a probability model for ***Z*** directly in terms of ***X***, UQ approaches model ***Z*** in terms of reality ***Y*** (and some error process) and then model ***Y*** in terms of the model state at a particular ‘best’ choice of input parameters, ***θ***^∗^. Letting ***Y***_0_ represent the elements of ***Y*** represented by the data ***Z***, the standard UQ-calibration model is

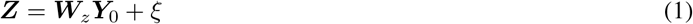

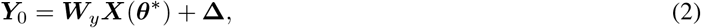

where ***W***_*z*_, ***W***_*y*_ are incidence matrices (e.g. ***W***_*z*_ might aggregate the represented model states, and ***Y***_0_ = ***W***_*y*_***Y***), *ξ* is an independent observation error with given distribution and **Δ** is model discrepancy, also with a given distribution and independent of ***X*** and ***θ***^∗^.

Acknowledging model discrepancy is critical to meaningful calibration of a model [Kennedy and O’Hagan, 2001, Plumlee, 2017]. Without model discrepancy, parameter estimates for ***θ***^∗^ will be biased, predictions will be both wrong and overconfident (with under-reported uncertainty), and hence any decisions made using calibrated predictions may be very bad (particularly if the under-reporting of uncertainty has led to underestimation of the risk of particularly bad outcomes). The parameters, ***θ***^∗^, and the discrepancy, **Δ**, are well known to be unidentifiable [Brynjarsdóttir and O’Hagan, 2014], meaning that in order for a calibration to be successful, detailed prior modelling of **Δ** (or ***θ***^∗^) is required.

There are two main approaches to calibration under model discrepancy: ‘Full Bayes’ [FB, Kennedy and O’Hagan, 2001] and History Matching [HM, Craig et al., 1996]. FB places a prior on the best input, *π*(***θ***^∗^) and a hierarchical Bayesian model on (1) to derive the posterior distribution *π*(***θ***^∗^ |***Z***) using, for example, Markov chain Monte Carlo (MCMC) methods [Gilks et al., 1996]. An example of the type of hierarchical model for counts within the general framework of (1) would be

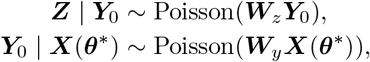

where the distribution of ***X***(***θ***) is represented by the ID model. In practice, the Poisson distribution here does not allow for a structured discrepancy model, so a better model might use a Negative Binomial distribution and use a careful prior for the dispersion parameter(s). HM requires only means and variances for each term in (1) and rules out values of ***θ*** when they lead to large values of a distance metric called *implausibility, ℐ* (***θ***), with typical form

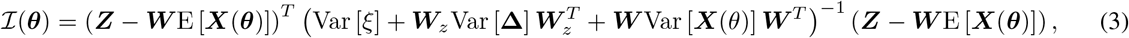

with ***W*** = ***W***_*z*_***W***_*y*_ [Salter et al., 2019]. *Note that if the ID model can be run for every evaluation of implausibility (3)*, *then the implausibility becomes*

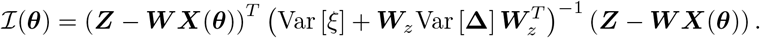

A threshold *T* is set and (3) is used to define a subset of parameter space, the Not Ruled Out Yet (NROY) space, of values of ***θ*** that may be consistent with (1). NROY space, **Θ**_*NROY*_, is defined via **Θ**_*NROY*_ = {***θ*** : ℐ (***θ***) ≤ *T*} . Structured discrepancy judgements come in the form of a prior elicitation of the covariance matrix, Var [**Δ**] [Vernon et al., 2010].

### 2.2 Emulation

Typically the ID model will be too computationally expensive to embed directly in MCMC (for FB) or to obtain a sufficiently large sample from **Θ**_*NROY*_ (for HM). Instead, an emulator is created to replace ***X***(***θ***) in either algorithm with a statistical model that is fast to evaluate. Whilst there are many different choices and variants of emulators that could be used [see Gramacy, 2020, for details on many of these], we focus our exposition on the most popular choice, the Gaussian Process (GP), and related models. For scalar model output, *f* (***θ***) say, a GP is denoted

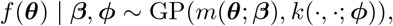

where the mean function, *m*(), depends on hyperparameters ***β*** and the kernel function, *k*(***θ, θ***^*′*^; ***ϕ***), depends on hyperparameters ***ϕ*** and represents the covariance, Cov [*f* (***θ***), *f* (***θ***^*′*^)]. The GP has the property that the joint distribution of *f* () at any finite collection of inputs, ***θ***_1_, …, ***θ***_*n*_, is multivariate Normal, with mean vector determined by *m*() and covariance matrix by *k*(). GPs have the attractive property that, having observed a training set ***F*** = [*f* (***θ***_1_), …, *f* (***θ***_*n*_)],

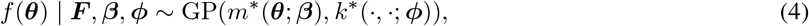

with known closed form expressions for *m*^∗^ and *k*^∗^. ‘Building an emulator’ then involves choosing the forms of *m*() and *k*(), and deciding how to ‘fit’ the hyperparameters, with options for the latter including maximum likelihood, cross validation, or, when embedded within MCMC for FB calibration, simply using Gibbs steps to update *π*(***β, ϕ***| ***F***) (certain choices of *m*() and priors also lead to being able to integrate out some of the hyperparameters analytically, Haylock and O’Hagan, 1996).

When calibrating with HM, you can use the GP or any emulator that gives a mean and variance. For ID models, Bayes linear emulators [Vernon et al., 2010] have often been used, which use the same specifications for *m*() and *k*() without the Gaussian assumption, yet have the same updated mean and covariance function, *m*^∗^() and *k*^∗^(), as in (4). Emulators have been extended to non-continuous and stochastic outputs, normally by specifying that the parameters of the data model (e.g. the variance of a continuous output, or the Poisson rate of a count) also follow a GP.

To calibrate with an emulator using FB, one must emulate ***W***_*y*_***X***(***θ***), which can be high-dimensional. For HM, an emulator is needed only for ***WX***(***θ***), which usually has fewer dimensions. Previous UQ approaches in ID modelling [for example Andrianakis et al., 2015, 2017] have looked to calibrate to high level aggregations of the data, meaning only a few emulators are needed (potentially for either approach). For example, Dunne et al. [2022] calibrate a COVID-19 model to total deaths from the whole of Scotland after 200 days of the pandemic using HM. This might be effective, particularly when the goal of the analysis is inference for the parameter vector ***θ*** (for example, learning *R*_0_), but may not be so effective when predictions at finer scales (e.g. regular 2 week forecasts at a local level) are needed.

Emulating and calibrating to high-dimensional data (such as spatio-temporal fields) can be achieved in a number of ways. A common approach is to fit independent GPs to every output [Johnson et al., 2015, Salmanidou et al., 2021], even when this leads to millions of GPs, and there exists bespoke software for this [Gu and Berger, 2016, Daub et al.]. There are emulation methods for time-series data [Mohammadi et al., 2019] and for joint emulation of multiple outputs [Conti and O’Hagan, 2010], yet by far the most popular approach is to project the high-dimensional output onto a low-dimensional basis, such as those derived from principal components, and to emulate the coefficients [Higdon et al., 2008, Salter and Williamson, 2022, Pepper et al., 2023].

Emulation in higher dimensions is challenging, more time consuming than for a single or handful of outputs, yet feasible. Salter et al. [2025] develops an emulator for the high spatial resolution version of the model we present in this paper and in Part 2. However, to calibrate in higher dimensions, model discrepancy over those dimensions is also required. Whilst the specification of high-dimensional structured discrepancy judgements is not infeasible *per se*, it is challenging and time consuming and has to be done for any new version of the model that might be used. In practice, that means that calibration using UQ takes time and requires a great deal of specialist input.

### 2.3 Critical challenges for real-time UQ in pandemics

#### 2.3.1 Seeding

ID models, ***X***(***θ***), require initial conditions in order to simulate the evolution of the disease, i.e. since ***X***_*t*_(***θ***) = ***X***_*t*_(***θ, X***_*t−*1_), then to run the model we must pass ***X***_0_ as well as ***θ***. The higher the spatial resolution of the model the more complicated the initial condition problem becomes. At the start of a pandemic, the traditional approach is to introduce a number of initial infected individuals into the system from the start (they can also be introduced gradually, though that adds to the dimensions of the initial condition problem). For brevity, we will refer to these initial infected individuals as “seeds”. Generally, we won’t know where the initial infections happened in reality, and so it is unlikely that the seeding we do use will be close to reality. It is worth noting explicitly that even if the best input, ***θ***^∗^, were known, realisations of ***X***(***θ***^∗^) may look nothing like the data if the seeding process is badly wrong.

If calibrating to high level summaries (like total deaths/infections) and if the goal of the analysis is only to infer the parameters ***θ***^∗^, it could be that the error introduced by using seeds is well captured by the discrepancy, **Δ**. When the number of infections begins to swamp the number of seeds, it’s also likely that the location and number of the seeds becomes less important for identifying the parameters. It may also be the case that forecasting the same high-level summaries shows sufficient skill over short time scales to help with decision support.

On the other hand, there are many situations where the initial conditions are critical either to the calibration or to the quality of the forecast or both. Consider, for example, the need to calibrate to spatial data in order to forecast the burden on individual hospitals for resource allocation. It may be that certain parameters control movements and hence the spatial dynamics of spread are needed to constrain them. It might be that the model is to be used to forecast the spread of a new variant, where the initial conditions include those recently recovered from a different variant and those that are vaccinated. Furthermore, unless a population is entirely closed, it will also be the case that new infections can be introduced at any time through mechanisms that may not be captured directly through the simulation model, such as unobserved movements of individuals across borders. When high dimensional data/forecasts are needed to support policy, these problems can only be overcome (within the traditional UQ frameworks described above) with a complex high-dimensional structured discrepancy model.

#### 2.3.2 Model evolution

ID models are constantly being adapted and developed in order to support policymakers as the situation evolves. A complex, accurate, high dimensional emulator and structured model discrepancy could take weeks or more to develop for a single model version. However, during the COVID-19 pandemic, a team would often only have a week from having been posed the latest policy support challenges to providing their forecasts to policymakers, including time to develop the model in order to answer the question [Brooks-Pollock et al., 2021]. As the pandemic evolves, different things must be modelled and different questions must be answered. Some examples from our work supporting the modellers who were informing UK policymakers during the COVID-19 pandemic included: an early focus on ICU bed occupancy and potential ventilator shortages; transmission within and between care homes; the importance of infections amongst asymptomatics; the potential efficacy of local lockdowns and ‘tiers’; vaccine efficacy; and waning immunity. The modellers must evolve their models constantly and adapt them to answer key questions. This might mean the introduction of new data streams and, in most cases, a completely fresh calibration/prediction task. The seeding problem compounds this difficulty and, unless the model is particularly fast to run and particularly simple to emulate, renders calibration/prediction using the UQ methods outlined above infeasible for supporting decision makers in real-time.

Typically, forecasts made using complex ID models will estimate parts of ***θ***^∗^ offline using other studies, not give a treatment to model discrepancy, and focus computational resource on providing forecasts that are consistent with ***Z***. The focus on data assimilation for short-term forecasting is both natural and important. Numerical Weather Prediction (NWP) takes a similar approach with great success [Hu et al., 2023], though model discrepancy is considered whilst parameters are fixed. However, although we believe that some form of data assimilation is critical, it is also the case that parameter uncertainty and model discrepancy are very important for ID forecasts, even on short timescales. It should also be noted that unlike for ID models during a pandemic, NWP models are rarely altered, allowing ‘pre-calibration’ and discrepancy estimation to happen during model development. In the next section we present a framework that combines the UQ ideas discussed above with data assimilation in a framework for real-time calibration of an ID model.

## 3 Data assimilation for ID models with embedded discrepancy

Our approach is to acknowledge that model discrepancy is present due to incorrect initial conditions and the simplified processes for transmission in an ID model, and to embed discrepancy in every element of the model state (*c*), at every age-class (*a*) and at every time point (*t*). Conceptually we consider the model

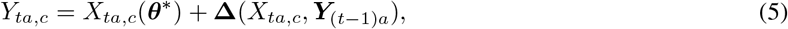

where we make the model discrepancy explicitly dependent on the current model state and the state of reality at the previous time. For ID models, the discrepancy depends on the model state because as *X*_*ta*,*c*_ increases, what we think of as an accurate prediction changes (we become more tolerant to errors). For example, suppose, early in a pandemic there is only one hospitalisation in the model state. When we think of the discrepancy as a process for correcting that count to the truth, we might specify something with a standard deviation of *±* 3 (say) cases, with anything more than that representing a poor model (note the actual choice here is model and modeller dependent). Later in the pandemic, if there are 1000 hospital cases, to have the same discrepancy uncertainty would claim that the model was almost perfect when, say, *±*50 might seem more reasonable (and still indicative of a skillful model). We develop the model discrepancy further in Section 3.1.

Suppose ***θ*** are fixed and the seeding denoted ***X***_0_, then a simulation of an ID model for *T* time steps is a draw from

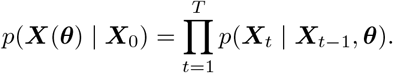

As discussed in the previous section, typical UQ approaches then look to obtain a collection of runs at different choices of ***θ, X***(***θ***_1_), …, ***X***(***θ***_*n*_), which are used for calibration once model discrepancy is added to (components of) ***X*** post-simulation. Embedding model discrepancy means, at each time-step of a simulation, sampling the discrepancy and working with the density of ***Y*** . Specifically, setting ***Y***_0_ = ***X***_0_ and assuming ***θ***^∗^ is known, a simulation would be a draw from

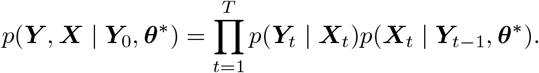

Suppose we have observations, ***Z***, for part of the state vector ***W***_*z*_***Y*** as discussed in Section 2, with a given time-independent observation error process, *p*(***Z***_*t*_ | ***Y***_*t*_). The likelihood, *p*(***Z***_1:*T*_ | ***θ***^∗^) is

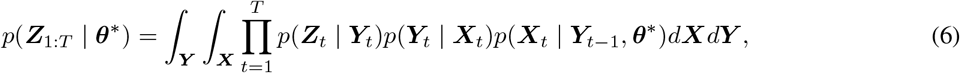

which cannot typically be evaluated analytically. Particle filters enable the likelihood to be estimated for any particular ***θ***, by setting off a number of particles (or ‘ensemble members’ in meteorological applications of data assimilation) that for each time point jointly sample ***X***_*t*_, ***Y***_*t*_ from the model, which are then re-sampled according to a weight determined by the data, before repeating this process for ***X***_*t*+1_, ***Y***_*t*+1_ and so on [see e.g. Doucet and Johansen, 2011]. By doing this, it is possible to generate unbiased and positive estimates of the likelihood that typically have a much smaller variance than other methods such as naïve Monte Carlo simulation. Furthermore, the particle filter can provide approximations to the filtering densities of the hidden states conditional on the observed data, which can greatly improve forecasting from any given model [Doucet and Johansen, 2011].

To fix ideas we describe a simple particle filter, the bootstrap filter [Gordon et al., 1993]. More complex variations for spatial ID models are developed in Part 2 [McKinley et al., 2025]. The bootstrap filter begins with *K* particles at ***Y***_0_ (or some draw from an initial distribution *p*(***Y***_0_)). At each time step, the *K* particles (with current values 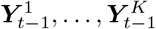) are evolved by sampling from the ID model, 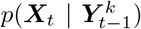, drawing 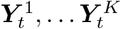 from the embedded discrepancy model, 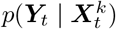, weighting each particle by 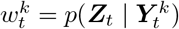, and then re-sampling *K* particles with replacement from the existing particles, 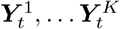, according to probabilities proportional to 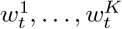. This process is shown in Algorithm 1. At each time step, we see the individual particles favoured are those closer to the data (hence the ‘data assimilation’), and the product of the weights over time averaged over the particles is an unbiased and positive estimator of (6).

It is possible to utilise these outputs in powerful ways. For example, Andrieu and Roberts [2009] showed that if an unbiased and positive estimate of the likelihood is used in place of the true likelihood in a standard Metropolis-Hastings algorithm, then the MCMC will converge to the true posterior distribution, *p*(***θ***^∗^ |***Z***_1:*T*_), in probability. This has facilitated the development of a suite of particle MCMC (pMCMC) methods that use particle filters to provide efficient estimators for analytically intractable likelihoods [Andrieu et al., 2010, Del Moral et al., 2015, Drovandi et al., 2016]. Though convergence to the posterior is assured, running the particle filter to evaluate (6) requires *K* evaluations of the ID model across *T* time points, where *K* is the number of particles. It will generally not be feasible to embed such a computationally expensive filter within an MCMC scheme for learning *p*(***θ***^∗^ |***Z***_1:*T*_), unless the ID model itself is inexpensive to run. On this point, many ID models are inexpensive, particularly those that are not spatial, or have low spatial resolution, and pMCMC has been applied to these before [e.g. McKinley et al., 2020]. If an emulator for the model was fitted and used to replace *p*(***X***_*t*_ |***Y***_*t −* 1_, ***θ***) in (6), that could make the calculation feasible for expensive models; however, emulation of the full model state vector is generally infeasible in these situations as discussed in Section 2.

### Algorithm 1

Bootstrap Particle Filter

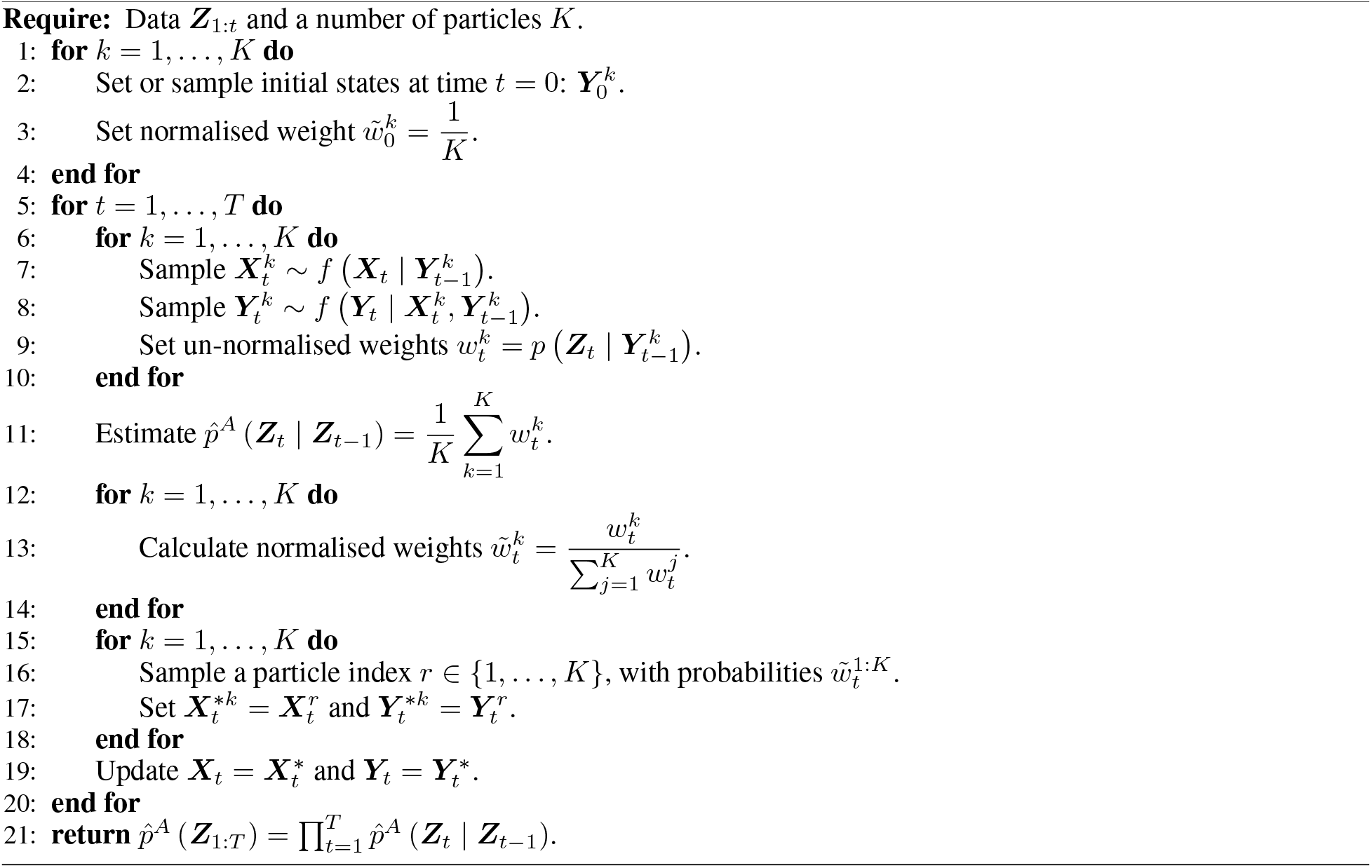

Our approach will be to emulate the likelihood based on a collection of ‘runs’ of the particle filter for a design ***θ***_1_, …, ***θ***_*n*_. Calibration using emulators of the model likelihood has been attempted before [see Oakley and Youngman, 2017, for example], and we present a particular method for using emulated likelihoods to do this in Section 3.2. What makes this idea particularly attractive for real-time calibration is that the emulation task is one-dimensional, no matter how complex and high-dimensional the ID model is. The emulation task is also independent of the complexity of the particle filter used to estimate the likelihood. Therefore, if the model can be developed to run in ‘data assimilation mode’, as is the case for climate models that are used in NWP, the UQ can be embedded relatively cheaply. Before discussing the emulation and calibration in detail, we present our method for embedding model discrepancy.

### 3.1 Embedded Model Discrepancy

A key source of dependence of **Δ**_*ta*,*c*_ on *X*_*ta*,*c*_ is that since the model state always represents counts that are bounded below by 0, **Δ**_*ta*,*c*_ must be bounded below by *− X*_*ta*,*c*_. A similar logical restriction on reality, *Y*_*ta*,*c*_, is the source of the dependence of **Δ** on ***Y***_(*t* − 1)*a*_. As the ID models we consider are compartmental models where, at any time, individuals can only move from their current state to a limited number of states, there is an implied limit on how many individuals can occupy a certain state that depends on the occupation of those states that lead there. Using the prime notation to denote the number of new entries into a given state (the *incidence*), then for example, an upper bound on new deaths in hospital at time *t* in the model introduced in Figure 1, 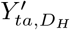, is the number of individuals in hospital at the previous time step, *Y*_(*t−*1)*a*,*H*_ .

Our discrepancy modelling for individual states makes use of a natural distribution for counting errors, the Skellam distribution (arising as the difference between two independent Poisson distributions). Letting random quantity *W* follow a Skellam distribution with parameters *λ*_1_ and *λ*_2_, the PMF is

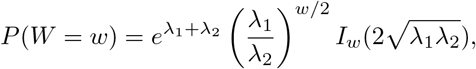

where *I*_*w*_(*x*) is the modified Bessel function of the first kind. As the probability mass function (PMF) of the Skellam is derived as the PMF of the difference between two Poisson distributions, the first with rate *λ*_1_ and the second with rate *λ*_2_, we have that

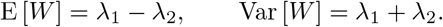

We can easily exploit these properties to make reasoned prior judgements. For example, setting *λ*_1_ = *λ*_2_ to ensure that the discrepancy has expectation 0, and selecting *λ*_1_ based on the size of the errors we might expect for a particular state of the model (as a hypothetical example, we might argue that a single transition of the model should be within 10 deaths of the true counts and to be outside that range would be a 2 standard deviation event, implying *λ*_1_ = 12.5). As our discrepancy model requires bounds, we truncate the Skellam both above, at *U*, and below, at *L*, and write this as

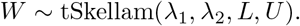

To make use of truncated Skellam distributions for modelling discrepancy, we factorise the discrepancy into the product of univariate densities that we model with truncated Skellam distributions and where the bounds can be derived from the conditional structure. The factorisation developed below is general and permits easy sampling from the discrepancy process as well as simple evaluation of the joint density for inference.

#### 3.1.1 Factorising the joint discrepancy

Noting that, if there are *N* model states, then there are *N −* 1 probabilistically valid factorisations of *p*(**Δ**_*t*_ | ***X***_*t*_, ***Y***_*t−* 1_) into the product of univariate conditional densities (we don’t need a discrepancy for *S* for a known population given all other discrepancies), the choice of factorisation we present here is a critical part of the proposed methodology for embedding discrepancy as it facilitates intuitive modelling of the conditional discrepancies. We present our factorisation in full generality, but will make use of the COVID-19 model presented in Figure 1 (introduced in Section 2) to fix ideas and motivate the methodology.

Let ℒ ^0^ be the set of absorbing states of the ID model, so that

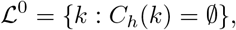

where *C*_*h*_(*k*) denotes the set of child states of state *k* (i.e. those states an individual can enter after arriving in *k*). The absorbing states are ordinarily the ‘recovered’ states and, in our example (see Figure 1) ℒ ^0^ = {*D*_*H*_, *R*_*H*_, *D*_*I*_, *R*_*I*_, *R*_*A*_} . For absorbing states, we apply our additive discrepancy model in equation (5) to the incidence counts, 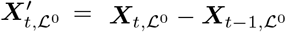, and we apply (5) to the raw counts for all other model states. The rationale for this difference is that it keeps the required subjective judgements (the mean and variance of the discrepancy) on similar scales throughout the model. We present the factorisation of the absorbing states only first, both as an important piece of the overall factorisation and to motivate our approach. Though the positioning of the states in ℒ ^0^ is arbitrary, it is assumed fixed throughout so that 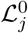 refers to the state in the *j*th position. The joint density of the discrepancy of the absorbing states at time *t* is

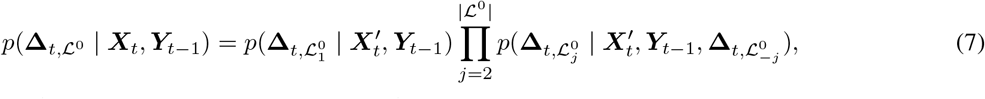

where *P*_*a*_(*k*) denotes the set of parent states of state *k*, and

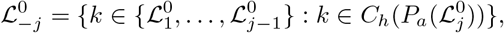

the set of absorbing states with position prior to *j* that share the same parents. Each of the PMF’s in (7) will be the PMF of a truncated Skellam distribution, where the bounds depend on the conditional variables. We present our choice for Skellam parameters (via specifying a mean and variance) in the next subsection, and motivate the above factorisation by deriving the bounds. For the COVID-19 model of Figure 1 (with the ordering of ℒ ^0^ as above), the factorisation in (7) is

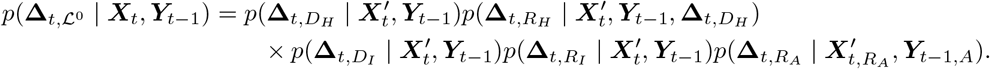

For discrepancies with no other discrepancy dependence, the bounds can be derived via (5) and via the idea that the states must be valid counts and that individuals entering a state at time *t* must have been in a parent state at time *t−* 1. For example,

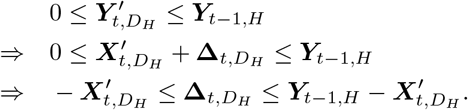

The dependency on other absorbing state discrepancies only occurs when absorbing states share a parent. Knowing the discrepancy for one state in these cases changes the bound as knowing the discrepancy implies a constraint on the available individuals in a parent node that can enter its other children. In the example, knowing 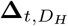 (and hence 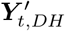) further constrains 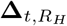 because

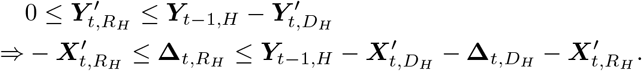

Let *L* represent the number of states in the longest pathway through the model (not including *S*). We will need to define a notion of stages in the disease pathway, with stage 0 being the absorbing states and then state 1 being the closest states connected directly to absorbing states and so on. Specifically, for *i* = 1, …, *L −* 1 define the set of states in stage *i*, ℒ^*i*^, where all child states belong to one of ℒ ^*j*^ for all *j < i* as

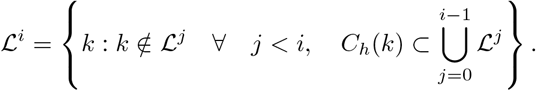

In our example,

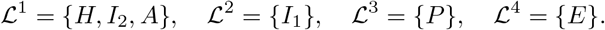

The full factorisation of **Δ**_*t*_ is

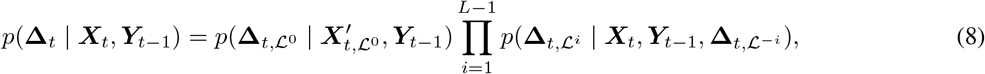

with 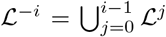 . Let *k* be a state in *ℒ* ^*i*^ and define the recursion 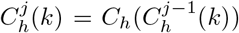 for 1 *≤ j ≤ i*, with 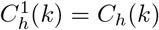 . The set of all descendants of state *k* ∈ *L*^*i*^, *D*_*e*_(*k*), is then 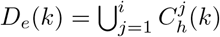 . For *j >* 1 define

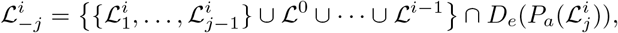

the set of states descending from the parents of 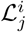 that also belong to ℒ ^*k*^ (*k < i*), or states in the current level in positions lower than *j*. Then, for *i* = 1, …, *L −* 1,

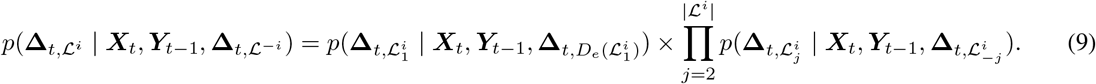

We give a more complex example of the implied bounds using this factorisation for the sets of non-absorbing states in (9) using our COVID-19 model in Figure 1. For 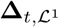, (9) gives

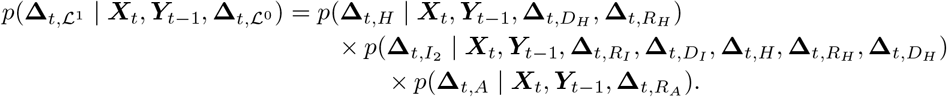

To see how the conditioning implies discrepancy bounds, take 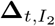, where we have that the incidence, 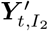, is bounded below by 0 (as usual) and by the number in *I*_1_ at *t −* 1 less those that moved into *H* or *D*_*I*_ at time *t*,

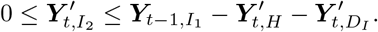

The hospital incidence, 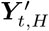, is the number entering *H* from *I*_1_,

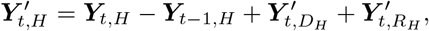

which is known given 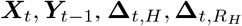 and 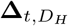 via

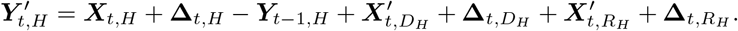

Therefore

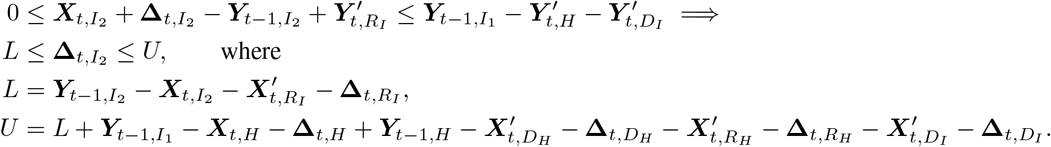

Whilst the factorisation of the discrepancy into sets via (8) is unique, the ordering of the states within each ℒ ^*i*^ is arbitrary and does alter the within set conditional distributions required. Our experience is that the chosen ordering is a matter of taste and does not impact the complexity of the bounds to be derived. Note that the factorisation method is fully general for any type of compartmental model structure, and the implied bounds are always derivable via simple rules that encode the dependence of bounds on eligible and previous moves between states. Full automation of factorisation and bounds for a particular modelling framework is therefore trivial, particularly if a random ordering is chosen within each ℒ ^*i*^, though modellers may prefer an ordering in terms of the severity of disease progression or something else.

#### 3.1.2 Discrepancy judgements

Whilst bounds for the individual time *t* conditional discrepancies can be trivially derived directly from the model structure as above, the other parameters of the Skellam distributions must be set and will determine the mean and variance of the (unbounded) discrepancy. The factorisation method developed in the previous section ensures that discrepancy can be considered carefully in terms of individual states at individual times at single locations if desirable, and it is important to be clear that any deeply held or carefully elicited judgements for the size of discrepancy can and should be used to determine these parameters where they exist. That said, whenever the model is altered (e.g. suppose a new state is added, such as an ‘Intensive Care’ state as a child of ‘Hospital’), we must be able to also make any alterations to the discrepancy at the same time in order to achieve real-time UQ that can be useful. We offer some principles that we have used to set the model discrepancy for our models here, and will revisit the topic in the discussion.

Our first principle is that *λ*_1_ = *λ*_2_ = *λ* for all states, times and locations. This choice states that though we accept discrepancy will be there, we don’t now have judgements about the direction of any specific model bias for a single time step (so given ***Y***_*t−* 1_). Our second principle is that the variance should be tied to the number of individuals in the state so that we allow for larger count discrepancies when the model count itself is already large. For our model and the examples in this paper, we set *λ* = *a* + *b****X***_*ta*,*c*_, so that the variance scales linearly with the counts and *a* and *b* are parameters to be chosen or estimated. Note that any increasing function of ***X***_*ta*,*c*_ would meet the given criteria, so, for example, we might have the standard deviation scale linearly with ***X*** or make some other choice.

The parameters *a* and *b* (or any other parameters of an increasing function of ***X***) need not be constant, particularly for different states or different locations. Our COVID-19 model does not have individuals from Scotland, so it might be that the discrepancy parameters are set to be larger at spatial locations in the North of England. We defer fitting the spatial version of our model to Part 2 [McKinley et al., 2025]. For this particular parameterisation, the *a* parameter plays the important role of controlling the model discrepancy when there are no cases. As discussed in the next section, we use model discrepancy to solve the seeding problem, by calibrating a data assimilation system where the model is always initialised without seeds. We return to particular choices of *a* and *b* in Section 4.2.

#### 3.1.3 Reporting errors

Completing the required statistical judgements for calibration requires specifying the properties of the observational error process, *ξ*, in equation (1). We noted in Section 2.1 that hierarchical models for counts might be more familiar in ID modelling, and, in fact, for the hierarchical Poisson model given there, *ξ* would follow a Skellam distribution (as the difference between two Poisson random quantities). Our model is

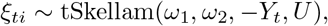

where upper bound, *U*, may be selected to avoid unphysically large errors and, as with our discrepancy model, the *ω*_*k*_ scale linearly with the number of cases:

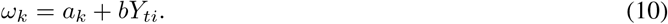

Here we don’t automatically fix *a*_1_ = *a*_2_, noting that, for example, setting *a*_1_ *< a*_2_ amounts to a judgement that on average the counts we observe in our datasets are lower than the true counts. For example, we might expect an under-reporting of deaths attributable to Covid when testing is not routine or robust at the beginning of a pandemic. As with our discrepancy judgements, linearity in the true counts is not necessary and is one possible choice that we make to minimise the number of parameters to elicit whilst giving some of the key behaviours that these errors should have.

### 3.2 Likelihood calibration

We now describe how we can use the embedded discrepancy model above with the data assimilation procedure in Section 3 to provide real-time calibration of an ID model so that the calibrated predictions can feed into policy support on the time-scales that they are needed, and so that the modellers have the freedom to adapt their model to answer key questions arising from a pandemic. Our method relies on being able to choose a collection of values of the parameters, ***θ***_1_, …, ***θ***_*n*_, and to run the model in ‘data assimilation mode’, by which we mean to run as a particle filter estimating (6) with embedded model discrepancy. Recall this means running *K* particles for each parameter choice ***θ***_*i*_ and at each time step being able to run the model from a discrepancy adjusted state (i.e. sampling from *p*(***X***_*t*_ |***Y***_*t −*1_, ***θ***_*i*_)). We note here that ‘data assimilation mode’ requires some software design and could not be set up in real-time for a brand new pandemic (any more than it is feasible to construct an infectious disease model from first principles immediately upon the emergence of a new pathogen). We are suggesting that model infrastructure work, which is done pre-pandemic, is rethought to allow an efficient data assimilation mode. Given this infrastructure, real-time changes to the model (e.g. the addition of compartments, connections or states, and the altering of spatial resolution), can be calibrated in real-time.

A major benefit of our approach is that we are able to set ***Y***_0_ = **0** at the start of the epidemic and allow the discrepancy model to add seeds into the system without needing to specify them. If we did this without discrepancy, the model would never have any infections. With model discrepancy, there is a small chance that infections are introduced by the discrepancy, and particles with introduced infections will be more likely to survive when the data support the presence of infection and more likely to be down-weighted and not re-sampled when there are no cases.

We work with the log-likelihood as this is numerically stable, though the particle filter estimate for the log of (6) is only asymptotically unbiased. We discuss the implications of this in our choice of filter for large models in Part 2. Let *l*(***θ***) = log(*p*(***Z***_1:*T*_ | ***θ***)) and 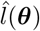 be an estimate of *l*(***θ***) made using a particle filter (for example the BPF described in Algorithm 1). Then, having chosen a design ***θ***_1_, …, ***θ***_*n*_, the ensemble 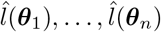 can be used to train an emulator using any of the methods described in Section 2.2. As the log-likelihood is scalar, emulation of it will usually be a relatively straightforward and automatable task for modern approaches (we say more about this as part of the examples in Section 4).

The likelihood calibration method introduced by Oakley and Youngman [2017] uses GP emulators for the log-likelihood and then an initial pre-calibration phase akin to history matching, where parameter values are ruled out if 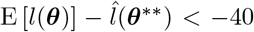, and ***θ***^∗∗^ denotes the location of the current observed maximum likelihood. They then have a method for adding runs to the ensemble until the emulator variance is almost 0 and can then use *e*^E[*l*(***θ***)]^ as an importance distribution for sampling from the posterior distribution. For the ID models we have worked with, though the likelihood emulation and pre-calibration steps work well, the variance due to the particle estimation was sufficiently large to ensure that *e*^E[*l*(***θ***)]^ was never a sufficiently good importance distribution.

The idea to rule out regions of parameter space where the likelihood is close to 0 underpins history matching [Williamson et al., 2015]. By explicitly emulating only the log-likelihood, we can implement a version of this idea directly. Though there are many ways we can consider doing this, a natural way is to attempt to rule out values of ***θ*** that are 1*/ϵ* times less likely than ***θ***^∗∗^ (with small *ϵ*). Using an emulator for the likelihood means that we need to factor in the uncertainty in *l*(***θ***) and we suggest that a natural way to do this is to define NROY space, **Θ**_*NROY*_, so that a point is ruled out if the probability that ***θ*** is 1*/ϵ* times less likely than our current best value exceeds *α*. More formally,

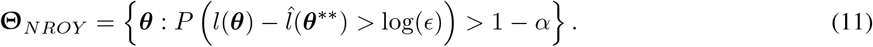

Note that either this probability is available directly from the emulator (in the case of a GP or other probabilistic surrogate), or can be accessed by a Cantelli inequality if using a second order emulator. In our examples, we set *ϵ* = 0.00001 and *α* = 0.95, though it may be that a higher/lower degree of certainty (higher/lower *α*) is required. The choice of *ϵ* defines the likelihoods that will be treated as 0 and this can be made smaller/larger in order to be less/more conservative in terms of ***θ*** retained.

Our approach is to mirror HM by performing a number of waves of likelihood calibration. For each new wave, running the particle filter for ***θ***_1_, …, ***θ***_*n*_ ∈ **Θ**_*NROY*_, fitting a new emulator using this ensemble and further reducing **Θ**_*NROY*_ according to (11) with this new emulator and where 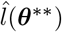 is the largest log-likelihood value seen through all of the waves. When the emulator variance no longer reduces significantly from one wave to the next, we can draw a final sample from **Θ**_*NROY*_ .

## 4 Application to an age-structured meta-population model for COVID-19

In this section we apply our framework to an age-structured meta-population model for COVID-19, first with a small population and using the model to generate data from which to calibrate, and subsequently for the whole UK in a simulation of the early outbreak. We first describe the model and choices that apply to both subsequent studies, before detailing each experiment.

The model for COVID-19, depicted in Figure 1, is a meta-population model where the individuals in each state transition (or not) to one of the subsequent paths in the network according to discrete probability distributions whose parameters are controlled by the model’s inputs. The model considers eight age-classes: under 5 year olds, 5–17 year olds, 18–29, 30–39, 40–49, 50–59, 60–69 and the over 70s.

Individuals in any age-class *a* move from *S* to *E* at time *t* with probability 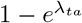, where

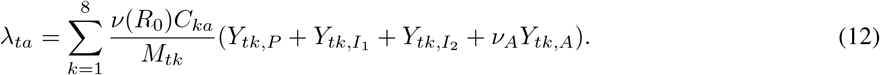

Here, *ν*(*R*_0_) is a deterministic function of *R*_0_ (see Supplementary Materials S2 for Part 2, McKinley et al., 2025), *C*_*ka*_ is the population contact rate for individuals in age-classes *k* and *a*, and *M*_*tk*_ is the number of individuals in age-class *k* at time *t*. We use the POLYMOD survey [Mossong et al., 2008] for our contact matrix prior to any lockdown and CoMix [Jarvis et al., 2020], thereafter. Effectively, this means that the number of individuals in infectious states within each age-class controls the probability of new infections along with the *R*_0_ parameter, and where only a proportion, *ν*_*A*_, of the asymptomatic state contributes to the infection rate. Our parameterisation only considers a single asymptomatic rate for the population, rather than age-dependent rates.

Once in the exposed state, *E*, and in subsequent states, individuals independently leave their state with probability 1 *−e*^*−γ C*^, where *γ*_*C*_ = 1*/T*_*C*_ and *T*_*C*_ is the average period spent in state *C*. 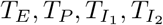 are calibratable parameters (see Table 1), 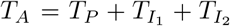, and time in hospital is modelled as a function of age (see below). Given an individual leaves their state at time *t*, they take one of the paths to a child node with probabilities modelled by age. Specifically, denoting the probability that an individual in age-class *a* transitions to state *C*^*′*^ from state *C*, as *p*_*a*,*CC*_*′*, we set

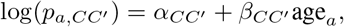

where age_*a*_ is the mid-point of age-class *a* and *α*_*CC*_*′* and *β*_*CC*_*′* are parameters to be calibrated. Specifically, for the intercepts we require 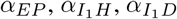 and *α*_*HD*_. We assume the same rate of change for each probability with age, setting all *β*_*CC*_*′* values to a single calibration parameter *η*.

**Table 1:**
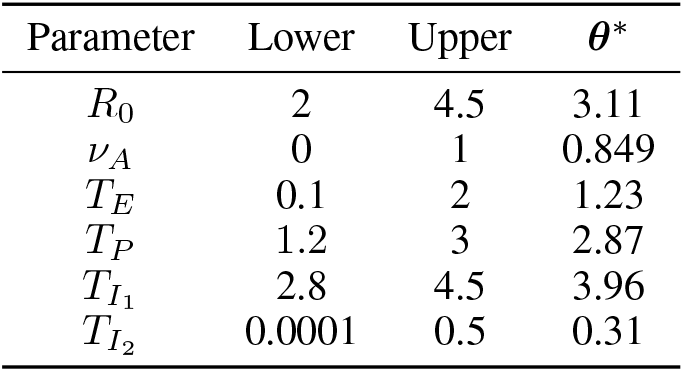
Parameter ranges and the values we chose for ***θ***^∗^ in our perfect model experiment. Parameters within the log-linear models are drawn from posterior samples of Bayesian regressions to external data as mentioned in the text and described in the Supplementary Materials Section S1.2.

| Parameter | Lower | Upper | $\theta^*$ |
| --- | --- | --- | --- |
| $R_0$ | 2 | 4.5 | 3.11 |
| $\nu_A$ | 0 | 1 | 0.849 |
| $T_E$ | 0.1 | 2 | 1.23 |
| $T_P$ | 1.2 | 3 | 2.87 |
| $T_{I_1}$ | 2.8 | 4.5 | 3.96 |
| $T_{I_2}$ | 0.0001 | 0.5 | 0.31 |

We also use a similar log-linear model for the mean hospital stay, *T*_*aH*_ (used for exit probabilities as described above), as a function of the median within-class age, 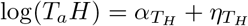 age_*a*_. These modelling choices were made at a time during the pandemic where log-linear models were already being fitted to describe age differences in seriousness of infection [Verity et al., 2020]. We say more about this in the Discussion (Section 6).

With the exception of the log-linear regression parameters, we elicited or derived parameter bounds with ID modellers on the project (co-authors Danon and Challen), or using data and the emerging literature during the pandemic. The ranges are given in Table 1, and a description of how they were arrived at described in the Supplementary Materials Section S1.2. For the *α* and *η* parameters, we used US data from the Centers for Disease Control and Prevention to fit relationships using Bayesian regressions. We fitted two models, one for 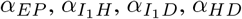 and *η*, and a second for 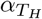 and 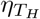 . For each model we drew a large number of samples from the joint posterior and then fitted a finite multivariate Gaussian mixture model (FMM) to these samples. This provided us with a tractable distribution around which we could build a bounded, uniform, but non-hypercube plausible region. The bounds of the plausible region were chosen by applying a threshold to the probability density function of the FMM, such that any parameter set with a probability density less than the threshold were considered outside the plausible region, and any with probability density greater than the threshold were considered inside the plausible region. The threshold was chosen by generating 1,000,000 samples from the FMM, and then taking the 0.001^th^ quantile of the corresponding 1,000,000 PDF values. Further constraints were placed to ensure valid probabilities for different transitions. For example, the log-linear model ensures that *p*_*a*,*CC*_*′ >* 0, but the prior space for the *α*_*CC*_*′* and *β*_*CC*_*′* parameters was truncated to ensure that *p*_*a*,*CC*_*′ <* 1 for all age-classes in the model. Similar constraints were also placed on any multivariate transitions, to ensure that not only was each probability bounded correctly, but also that the sum of probabilities equalled 1. Any parameter sets that violated these conditions were excluded.

Since we wanted the plausible region to be uniform within these bounds and constraints, we generated initial candidate values by sampling a large set of samples from the fitted FMM, excluded any invalid values, and then subsampled the remaining ones using a space-filling maximin design. The US data and the posterior sampling of our regression models is described in the Supplementary Materials Section S1.2.

### 4.1 Discrepancy and observation errors

We use the factorisation of the joint embedded discrepancy as presented in Section 3.1.1, with the ordering for each ℒ^*k*^ as given in our illustration. Namely,

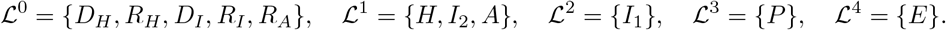

We use the principle for setting discrepancy judgements, *λ* = *a* + *bX*_*ti*_, described in Section 3.1.2. We choose to fix *a* and *b*, though note that, with a sufficiently strong prior, these parameters could be made calibratable. Given that we initialise without seeds, the *a* parameter determines the chance of introducing a case through the discrepancy. We want this to be small; otherwise we will quickly be simulating very large, discrepancy driven outbreaks. A simulation study showed that fixing *a* = 0.05 gives approximately a 95% chance (under a range of upper bounds) of 0 individuals being drawn from a Skellam distribution with no prior cases. Lowering *a* would reduce this further, which might be a reasonable thing to do yet, unless a very large number of particles can be generated, it may be difficult to get particles that trigger an outbreak. We choose *b* = 0.01 trading off a desire to keep the probabilities of extra cases low for low numbers of cases, with a desire for this probability to increase with *X*. This choice leads to a roughly 20% chance of more than 5 cases added in the discrepancy when *X* is 1000, which may be slightly low. Alternative non-linear forms of discrepancy function may be useful in growing the discrepancy more than this as cases increase, though in our spatial implementation of the model, the cases in each spatial unit are sufficiently small for this not to be an issue (see Part 2, McKinley et al., 2025).

For the reporting error process described in Section 3.1.3, we set the parameters in equation (10) as *a*_1_ = 0.01, *a*_2_ = 0.2 and *b* = 0.1. With these choices, there is a 20% chance that if there is a single case, it won’t be reported. For large numbers of cases the chance that the truth is under reported rises from 30% (3 cases) to 50% (1000 cases). We discuss inferring the parameters of the observation error and the discrepancy further in Section 6.

For moderate *λ*_1_, *λ*_2_ and large upper bounds, sampling the truncated Skellam within a particle filter can prove computationally demanding. This is due to the fact that the acceptance rates for a rejection sampler handling the truncation can become low for some particles as *λ* (and hence the variance) increases, and to the fact that the inverse transform for rejection-free sampling requires evaluating the mass function across the large population. Noting the Normal approximation to the Poisson distribution for large enough *λ*, we explored the efficacy of Normal approximations to the Skellam distribution and found very good accuracy for *λ >* 10. For discrepancy, we therefore sample from discrete truncated Gaussian distributions if *λ >* 10. We describe the sampling for this in the Supplementary Materials Section S1.

We implement the bootstrap particle filter described in Algorithm 1 with 500 particles in all of our experiments (described below). Each collection of particles is used to calculate the log-likelihood in (6). The resulting log-likelihood estimates are used as training data for deep Gaussian process (DGP) emulators [Ming et al., 2023] with heteroskedastic likelihoods (to account for the variable nature of the uncertainty on the log-likelihood as we move through parameter space). These are implemented in the dgpsi R package [Ming and Williamson, 2024], with a tutorial on implementing the specific heteroskedastic DGP available at https://mingdeyu.github.io/dgpsi-R/articles/motorcycle.html. We used DGPs as we found the log likelihood surface was highly non-stationary. These emulators are used to approximate the NROY sets given by (11), with *α* = 0.95 and *ϵ* = 10^*−*5^. NROY simulations from previous waves are included in the training so that, for later waves with the perfect model we had ensemble sizes approaching 1500. It is possible to train DGPs with dgpsi using the Vecchia approximation for large ensembles such as these and return an answer in a few minutes. Each wave took around 6 minutes for the particle filter to complete and a few minutes (depending on ensemble size) for the DGP, meaning that 10 waves could easily run in a couple of hours on a standard laptop computer.

### 4.2 Perfect model experiments

To demonstrate the method, we first simulate an outbreak within a small population of 10, 000, generating data, ***Z***, directly from equations (5) and (1) using the parameter values ***θ***^∗^ given in Table 1. We distribute our population across the age-classes according to the proportions in the true population (England), so that, from youngest age-class to oldest, the population sizes are 600, 1540, 1540, 1340, 1280, 1340, 1050, and 1310. To demonstrate the inference our model performs without seeding the outbreak artificially (letting the discrepancy enable the introduction of infection through the particle filter), we begin with all individuals in the *S* state and perform simulations of 60 day outbreaks. We allowed for 80% of the simulation time to represent the ongoing outbreak (for which we have data) and for the goal to forecast the final 12 days. We do a first experiment assimilating only deaths and a second assimilating both deaths and hospitalisations. Both experiments perform 48 days of assimilation before allowing all particles to continue their trajectories for 12 days.

We conducted nine waves of likelihood calibration as described in Section 3.2, each wave using 250 uniformly sampled points from the previous NROY set (with Wave 1 points chosen via maximin Latin Hypercube), with 200 points used to train DGPs and 50 design points for out-of-sample validation checks (following standard 80 : 20 ratios for model training and validation samples). A 10^th^ ensemble is then run with the particle filter from the Wave 9 NROY set to be used for inference and prediction. Samples for each wave are generated using an efficient slice sampler developed for history matching [Andrianakis et al., 2017], available within the hmer package [Iskauskas et al., 2024].

#### 4.2.1 Assimilating death counts only

Following all waves 84% of parameter space has been removed and the volume of NROY has converged (see column 2 of Table 2). This is not a large amount of space reduction relative to the later experiment and to other HM studies, pointing to the fact that deaths-only data has a limited ability to discriminate between parameter settings. Figure 2 plots the samples from NROY space for each of the 10 waves. The plots on the diagonal show the density of the samples in each wave, and the plots in the lower triangle of the array are 2D scatter plots coloured by wave number. The true parameter values, the ***θ***^∗^ from Table 1, are shown as red points. From this plot, we see that most of the constraint on the parameter space has come through constraining 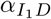, the intercept on the log-linear model for death rates in the community. The lowest and highest coefficients of age in the log-linear models, *η*, are constrained as the calibration moves through the waves; however, the other key parameter controlling death rate, *α*_*HD*_, is only marginally constrained. We believe that this is because either we can have a low death rate in hospitals with a large number of hospitalisations, or a high death rate with low hospitalisations and the data, as assimilated, is not able to distinguish between the two. Similarly, we don’t see a lot of constraint on parameters like *R*_0_, as the deaths we do see might be explained by high levels of disease and low death rates, or high death rates but low levels of infection.

**Table 2:** Proportion of original parameter space remaining after each wave of likelihood calibration rounded to 3 significant figures, for our 2 perfect model experiments and the calibration to real COVID-19 data presented in Section 5.

| Wave | Deaths Only | Deaths + Hospital Cases | Real Data |
| --- | --- | --- | --- |
| 1 | 0.287 | 0.560 | $8.10 \times 10^{-2}$ |
| 2 | 0.250 | 0.318 | $8.42 \times 10^{-3}$ |
| 3 | 0.244 | 0.272 | $2.95 \times 10^{-3}$ |
| 4 | 0.206 | 0.220 | $1.18 \times 10^{-5}$ |
| 5 | 0.180 | 0.201 | $7.08 \times 10^{-8}$ |
| 6 | 0.173 | 0.189 | $3.54 \times 10^{-10}$ |
| 7 | 0.160 | 0.180 | $7.08 \times 10^{-12}$ |
| 8 | 0.160 | 0.173 | $3.89 \times 10^{-13}$ |
| 9 | 0.160 | 0.162 | $1.68 \times 10^{-14}$ |

**Table 3:** Prediction intervals and median projected hospitalisations and deaths for all 4 lockdown scenarios 2 weeks after lockdown.

| Metric | Scenario | PI (50%) | PI (90%) | Median |
| --- | --- | --- | --- | --- |
| Deaths | No Lockdown | 7,254–37,268 | 3,138–192,138 | 15,171 |
|  | Lockdown Light | 1,272–46,648 | 489–5,615 | 2,698 |
|  | Lockdown Half | 1,189–4,601 | 417–33,104 | 2,032 |
|  | Lockdown Severe | 1,373–4,626 | 501–27,877 | 2,385 |
| Hospitalisations | No Lockdown | 27,030–117,516 | 11,463–658,269 | 52,512 |
|  | Lockdown Light | 3,543–15,513 | 1,428–156,081 | 7,532 |
|  | Lockdown Half | 3,263–12,297 | 1,296–98,643 | 5,337 |
|  | Lockdown Severe | 3,931–12,084 | 1,644–81,064 | 6,101 |

**Table 4:**
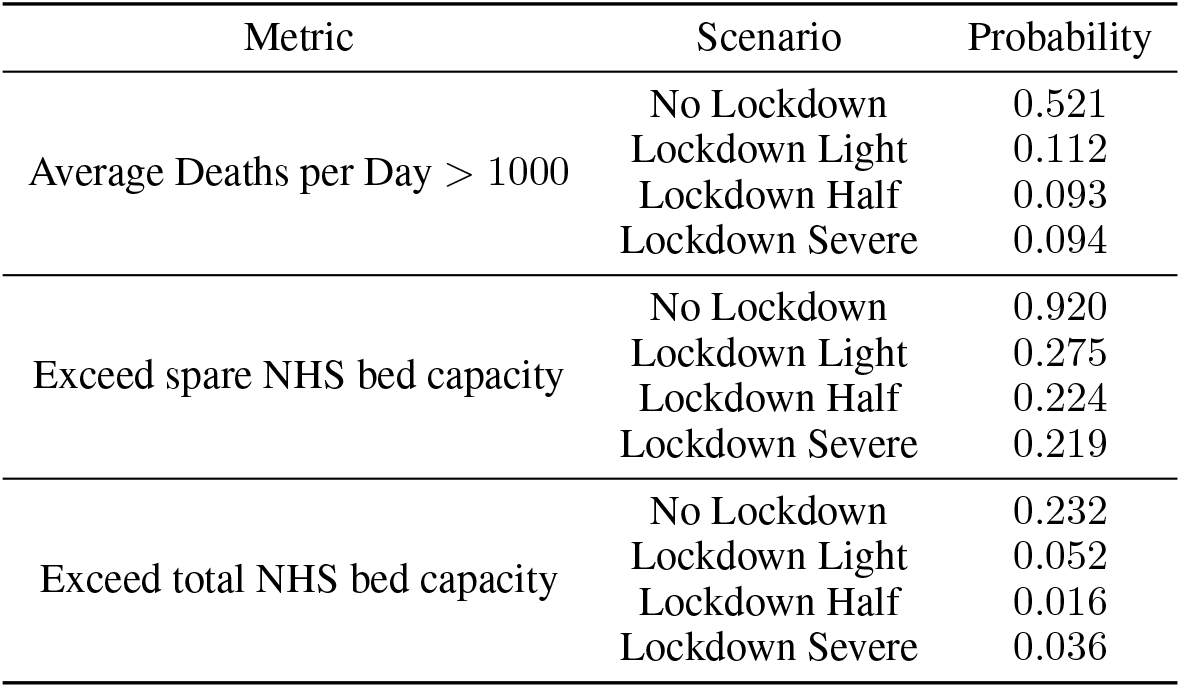
Summary statistics for policymakers for the four different lockdown scenarios.

**Figure 2:**
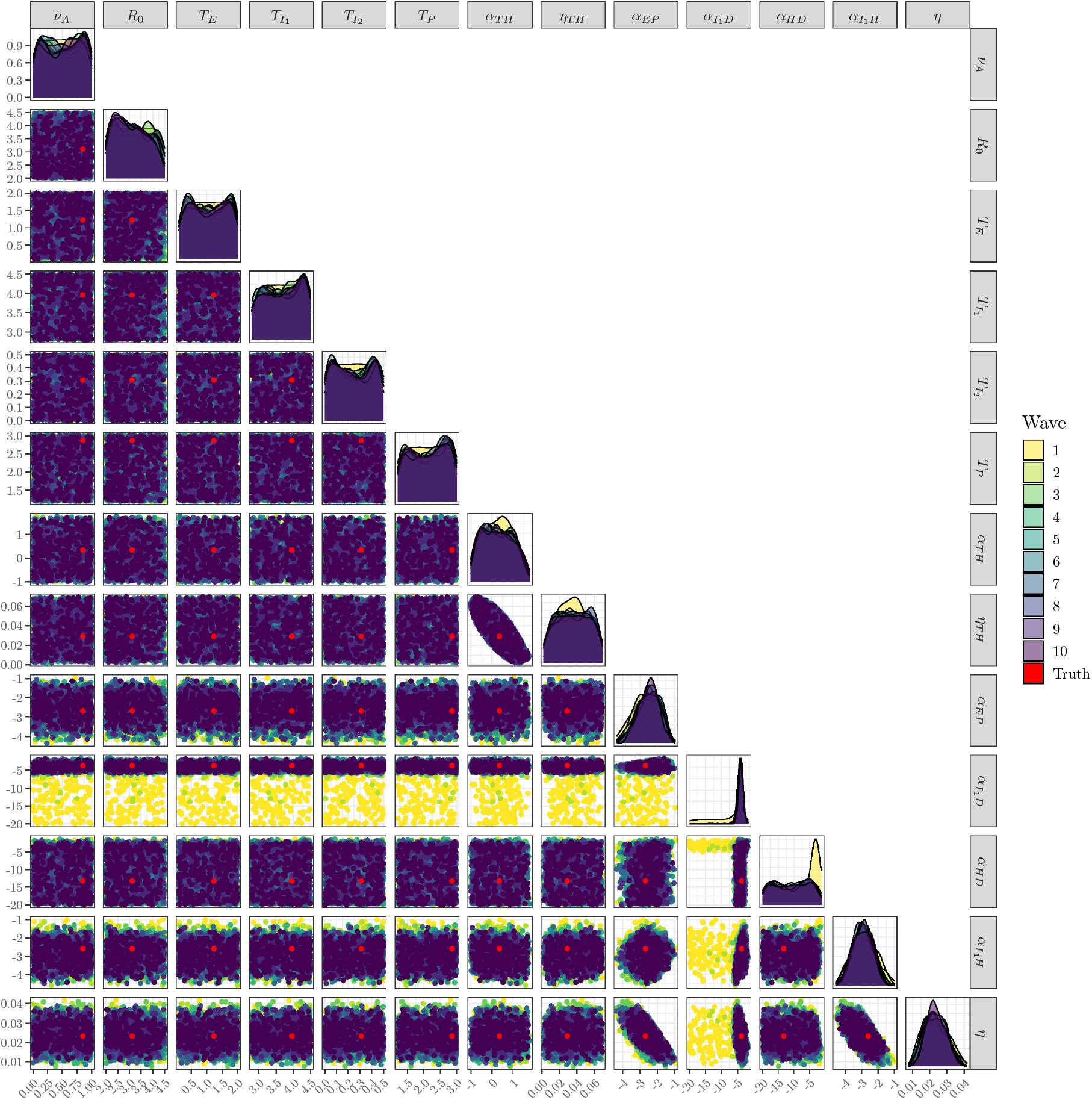
NROY samples for each of 10 waves for the perfect model experiment calibrating to deaths counts only. 2D scatter plots of NROY samples are plotted in the lower triangle of the array and are coloured by wave. The images on the diagonal are 1D density estimates calculated from the samples.

Figure 3 shows the particle trajectories for the wave 10 simulations for all states and age-classes in the model. As this is a perfect model experiment, we know all of the true hidden states, and we plot these in red. The dark blue dashed lines are the data we calibrate to (here the sum of *D*_*H*_ and *D*_*I*_), and we shade the region spanned by the 50% credible interval of the estimated states in darker blue and the 95% interval in light blue. The median simulation is given a solid line, whilst we indicate the end of the assimilation period with a vertical dotted line. From this plot we see that the particle filter tracks the data well and we have estimated the true disease states well. We note that, for this particular setting of the parameters, the disease has already passed its peak in our small population.

**Figure 3:**
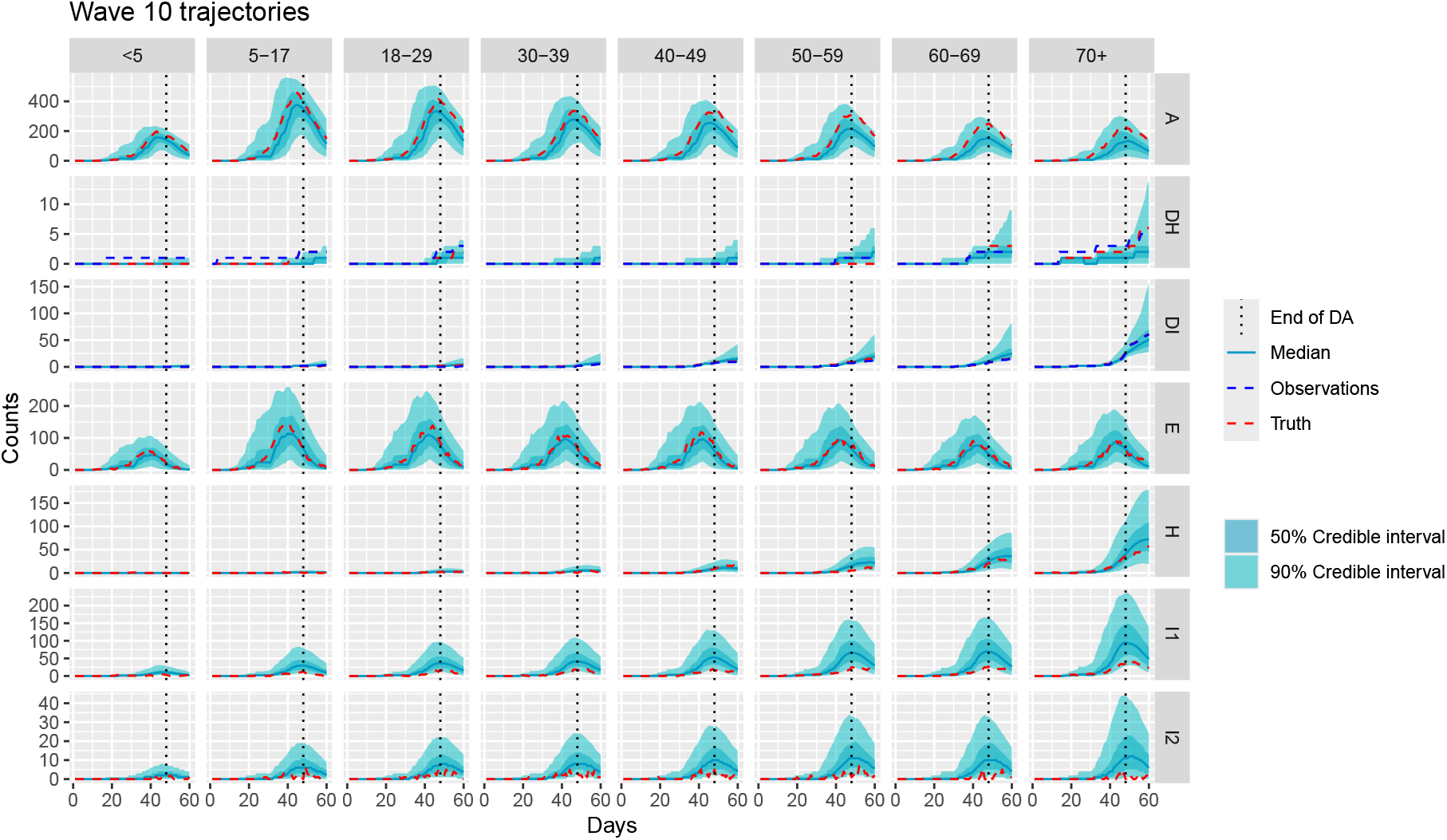
Particle trajectories for the wave 10 simulations for the perfect model experiment calibrating to deaths only. The true hidden states are plotted in red. The dark blue dashed lines are the data we calibrate to and we shade the region spanned by the 50% credible interval of the estimated states in darker blue and the 95% interval in light blue. The median simulation is given a solid line. The end of the assimilation period is represented by the vertical dotted line.

#### 4.2.2 Assimilating death counts and hospital cases

We repeat the history matching exercise above with the same perfect-model data as with the deaths-only experiment, and include hospital cases within the assimilation steps. After 9 waves we had removed 84% of the initial parameter space (column 3 of Table 2). We note that we ruled out less space by including hospitalisations and that we have not yet converged to a final NROY size by the end of the 9 waves. In standard history matching, adding metrics can never increase the size of NROY; however, here we are calibrating to the likelihood and new observation types can fundamentally change the likelihood surface. We say more about this in the discussion of Section 6. Figure 4 shows 10 waves’ worth of NROY spaces. In contrast to calibrating to Deaths only, we have ruled out more of the high *R*_0_ and low exposed waiting times, *T*_*E*_.

**Figure 4:**
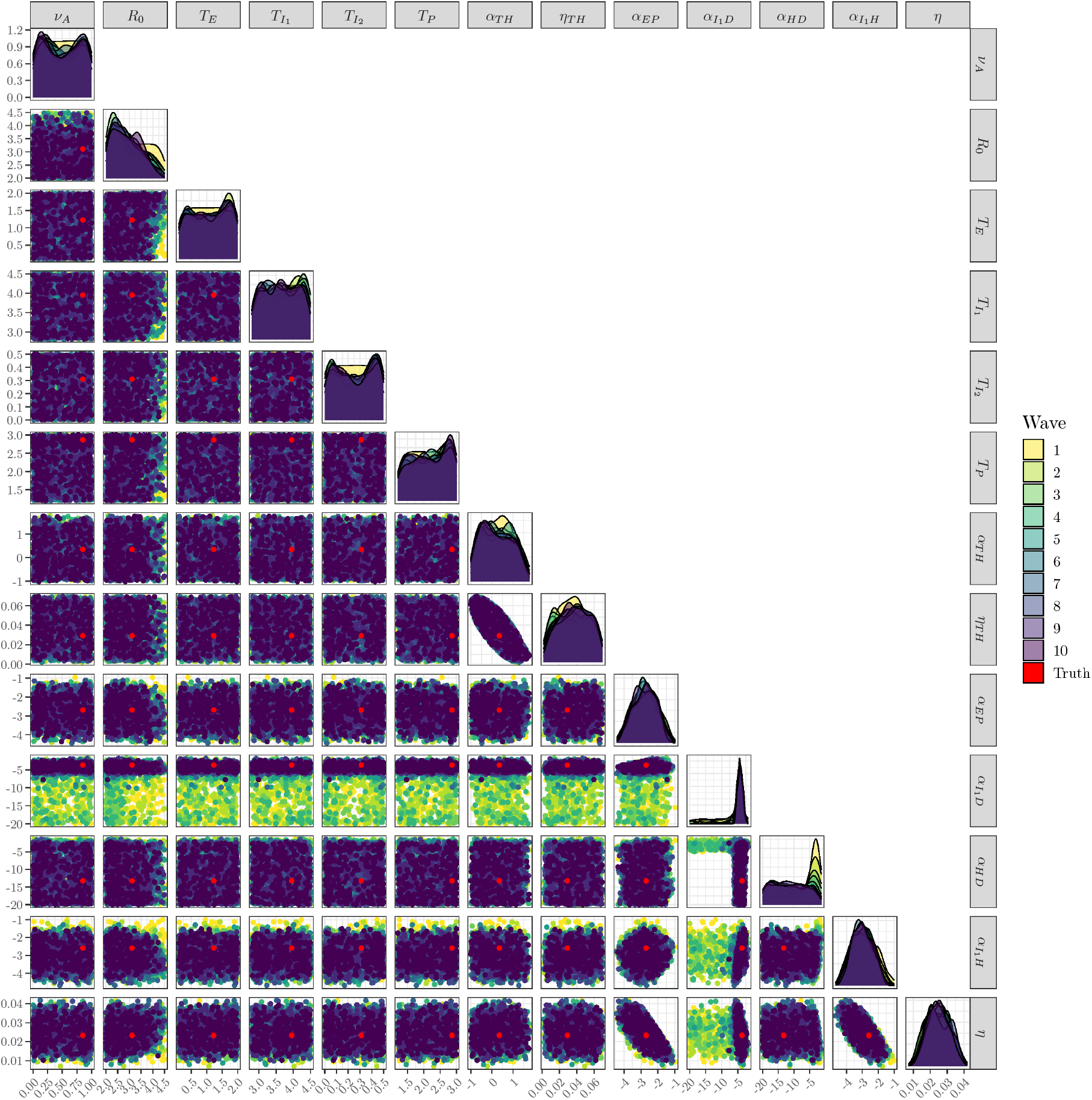
NROY samples for each of 10 waves for the perfect model experiment calibrating to deaths counts and hospital cases.

Figure 5 shows the model trajectories with the hospital state now having a blue-dashed line for observations (in contrast to Figure 3 where only the death counts are observed). Here we see good agreement with the data, yet some features of the true state (e.g. asymptomatic rates) are under-estimated. It may be that running more waves would rectify this issue; however, to ensure our experiments were automatable, we fixed the number of waves at 10. We discuss this further in Section 6.

**Figure 5:**
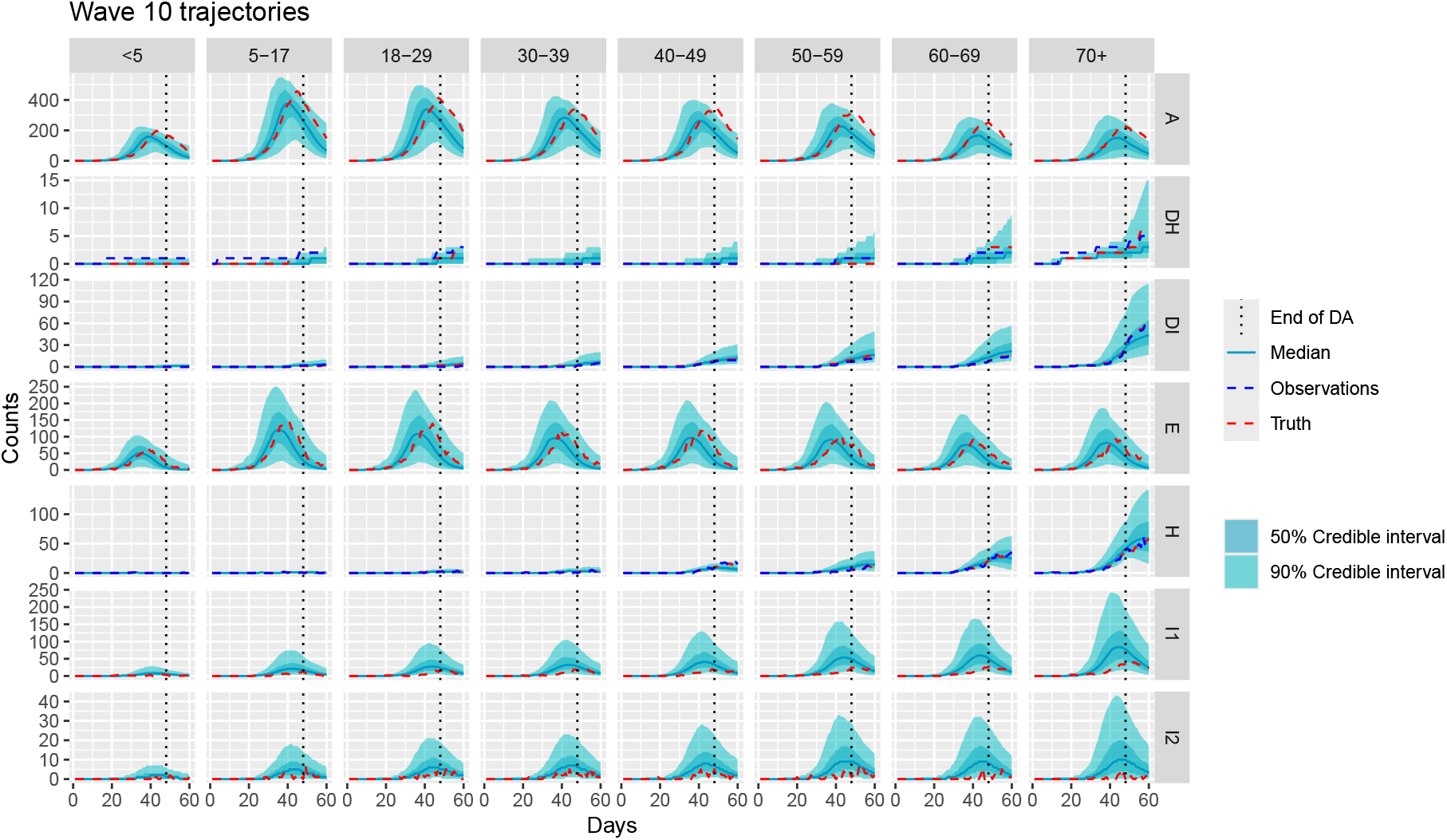
The particle trajectories for the wave 10 simulations for the perfect model experiment calibrating to deaths and hospital cases.

## 5 Application: UK COVID-19 lockdown, March 2020

We present a retrospective application to supporting policymakers during the beginning of the COVID-19 pandemic in the UK. At that time, modelling groups participated in the SPI-M-O subgroup of SAGE. Model results and uncertainty analyses were often synthesised prior to reporting to government [Silk et al., 2022, Bowman, 2022], and the turnaround time for answering questions was under one week (with the time with the latest data to support the answer around one day). We run our analyses as though changes to the model and amendments to the discrepancy structure are made during the week prior to the data arriving and as though our UQ must run overnight. Our population is the same size as the England and Wales population (at that time) separated into age-classes according to the true population proportions. The question we attempt to provide uncertainty analysis with our model for is the question of how effective would ‘lockdown’ be. Lockdown represented a severe restriction of movements exempting only a few key worker types and children of those key workers from travel, and a restriction of recreation activities to a single hour of solo outdoor activity for exercise.

We model restriction of movements using a scaling, *β*_*s*_, on the force of infection, *λ*, in equation (12) and by restricting the intra-generational mixing via the contact matrix. To simplify the narrative, we use the CoMix contact matrix [Jarvis et al., 2020] here, but if this analysis were undertaken prior to the policy decision, we would have needed to apply scalings to the contact matrix by hand. Note that our *β*_*s*_ and any intra-generational mixing parameters would, in principle, be calibratable after lockdown had happened, but at this point in the analysis we can only fix these and simulate forwards. Our goal is to predict with uncertainty, but to do this fully probabilistically, including an integration over the uncertainty in the *β*_*s*_ and the contact matrix may be unrealistic or hard to interpret for policymakers (or those performing synthesis). A different option is to analyse a handful of “scenarios” (corresponding to fixed values of the *β*_*s*_) and to give probabilistic uncertainty for these that includes discrepancy and parameter uncertainty. It would not be unusual for SPI-M-O to agree the nature of the scenarios to report on to enable meaningful synthesis across models.

We present a scenario analysis offering calibrated UQ of business as usual (no lockdown) and 3 types of lockdown, one that reduced force of infection by 20% (lockdown ‘light’), one by 50% (lockdown ‘half’) and one by 80% (lockdown ‘severe’). We note that putting a prior on *β*_*s*_ ∈ [0, 1] and integrating it out in the uncertainty analysis is as easy within our framework as performing these fixed scenarios, so there is nothing methodologically precluding it; however, it would be very difficult to disentangle the prior from the UQ (some of which speaks to the information in the data about the true state of the pandemic). We also note that, if a scenario had been prescribed to us, we might propagate uncertainty (on the *β*_*s*_, say) to represent uncertainty in that specific scenario. For example, we might consider ‘severe’ restrictions represented by *β*_*s*_ ∼ *β*(1, 4), which has prior mean 0.2. Our approach readily enables this uncertainty to be propagated through the trajectories.

### 5.1 Data

We calibrate to deaths by age group and total hospitalisations across all age groups during the assimilation phase of our model runs. We are not able to calibrate age-structured hospitalisations (according to our defined age structure) as NHS age-classes are different to the age-classes we have for deaths. We propose a solution to this in our spatial implementation in Part 2 [McKinley et al., 2025]. Our data assimilation period starts from 15^th^ January 2020, up to the 23^rd^ March 2020, the date of the first lockdown. Lockdown was on a Sunday, yet for reporting and synthesis it is possible that analyses such as these would be performed earlier in the week (say based on Tuesday’s data). For a decision as impactful to people’s lives as lockdown, it is likely that the most up to date data/forecasts might have been considered, or might be in a future pandemic, so the ability to turn around a re-calibration to whatever new data was available in a short amount of time might allow for data all the way to lockdown to be accounted for (in the projections).

We use deaths reported within 28 days of a positive test and obtained from https://coronavirus.data.gov.uk/. As the record begins on 2nd March 2020, we assume that deaths from COVID-19 are zero between 15th January–2nd March. The daily hospitalisations for this period are not available publicly and were obtained from the COVID-19 Hospitalisation in England Surveillance System (CHESS).

### 5.2 Calibration results

Figure 6 shows the NROY space after 10 waves of calibration to the observations described above. It is striking that, unlike for the perfect model experiments, we have converged to a specific region in parameter space and so are able to make specific inferences about the nature of some of the disease parameters. Before addressing these at face value, it is worth noting that the extent to which this inference is trusted should, as ever, represent the result of careful discussions with the modeller who will ultimately report them. Here, much of the inference is believable (and we’ll focus on these elements next), but certain things may be of concern. For example, the reduction in the force-of-infection due to asymptomatic cases, *ν*_*A*_, is surprisingly precise given that we have no data on asymptomatic cases. We believe the cause to be two-fold. Firstly, the data do speak to *η*, showing it to be high and implying older people are much more likely to suffer serious illness, hospitalisation and death from COVID-19. The prior on *α*_*EP*_ then forces the intercept on the log probability of moving into the symptomatic state, *P*, rather than *A*, to be in the lower half of its range (see bottom row, fifth panel from the right of Figure 6). That means a lower probability of symptomatic infection amongst the younger age groups and therefore higher probability that they enter the asymptomatic state. Given the inter-generational mixing and the hospital cases and deaths we do see, it can’t be that the force of infection is 0, nor as high as for the symptomatic cases.

**Figure 6:**
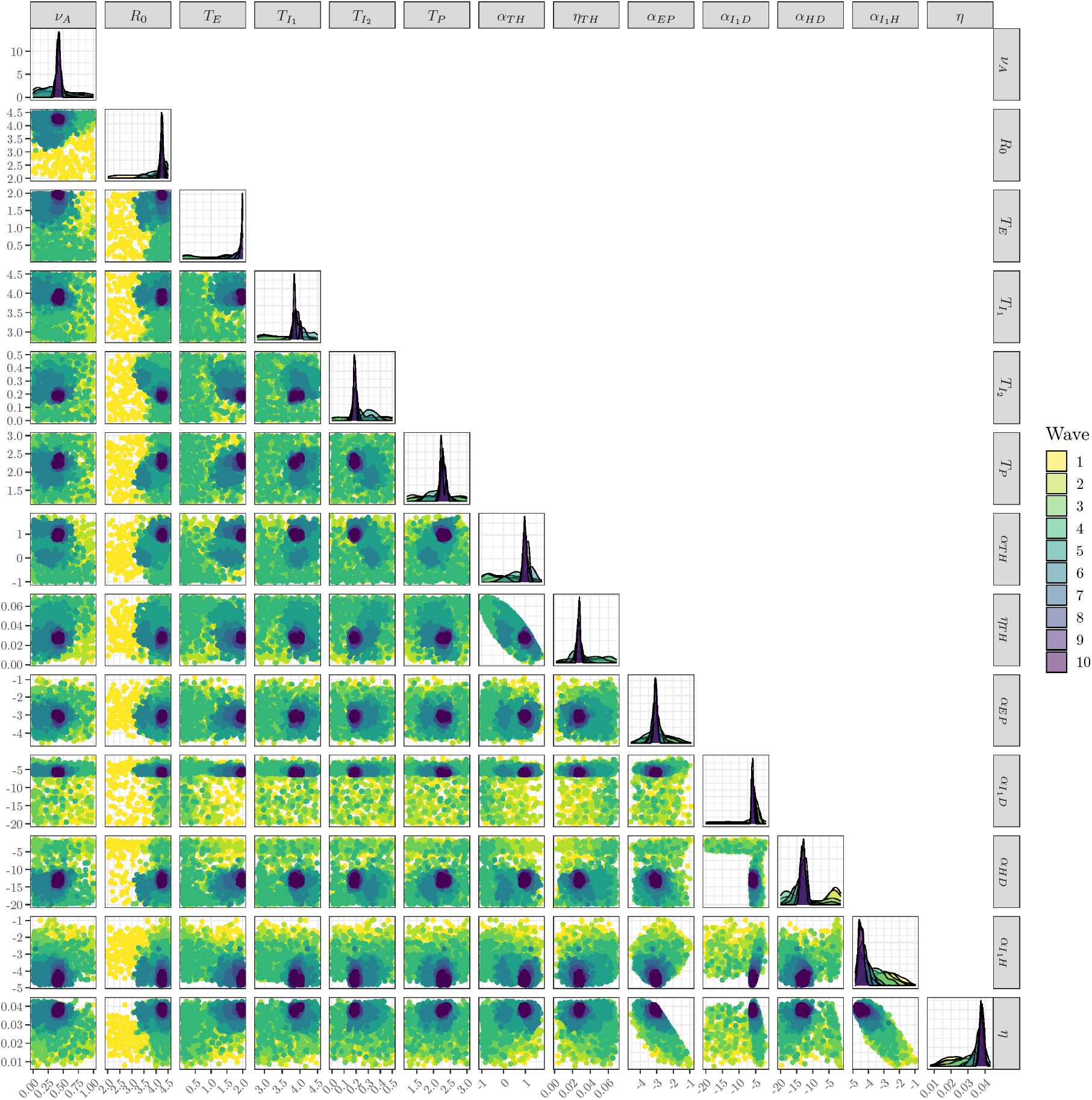
NROY samples for each of 10 waves for the calibration to age structured observations of deaths and hospitalisations up to lockdown. 2D scatter plots of NROY samples are plotted in the lower triangle of the array and are coloured by wave. The images on the diagonal are 1D density estimates calculated from the samples. (a) Trajectories of total cumulative deaths and hospitalisations across all ages after 10 waves. The left panels show the uncertainty in the projections up to 2 weeks after lockdown under the ‘severe’ lockdown scenario represented by 50% and 90% prediction intervals. The right panels zoom in so that the quality of the calibration to data prior to lockdown can be assessed. (b) The age-group trajectories after 10 waves of likelihood calibration for the ‘severe’ lockdown scenario.

There is, perhaps, another effect that is due to the use of the bootstrap particle filter rather than more advanced filtering techniques. As we start with no seeds and we begin seeing hospitalisations and deaths, which are at the end of the disease pathway, the BPF, as it only considers the current state, will not begin favouring introducing seeds via the model discrepancy until it sees its first cases in hospital. By then, in the data, there is infection around and it may have been around for some time. It can be that higher *R*_0_ values can compensate for introducing infections late, quickly bringing lots of cases into the disease states in order to get enough so that there are some that move quickly through to the hospital and death states. Given the size of the populations in each state and the amount of time our filter is assimilating data, this might not be a problem here, but can be a problem in spatial implementations as we will discuss in Part 2 [McKinley et al., 2025].

We also see that the data favour a higher baseline rate for deaths in the community, 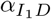, but a mid-range baseline for deaths in hospital, *α*_*HD*_. There is also a low baseline for hospitalisations from the *I*_1_ state (shown via 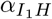). The parameters favour longer hospital mean waiting times through the larger value of *α*_*TH*_ . The large number of cases implied by a high *R*_0_ appears to be offset by a long mean waiting time of two days in the exposed but not yet infectious state, *T*_*E*_.

Figure 7 (a) shows the wave 10 particles for cumulative deaths and hospitalisations including the trajectory we have from the data. We show the full trajectories in the left panels and zoom in to the scale of the observations by lockdown (the vertical dotted line), plotting the data we assimilated (blue dashed line) against intervals calculated from the particles. The post-lockdown trajectories shown are those for the “severe lockdown”, where the force of infection is reduced by 80%. We discuss these and the projections for the other scenarios in Section 5.3.

**Figure 7:**
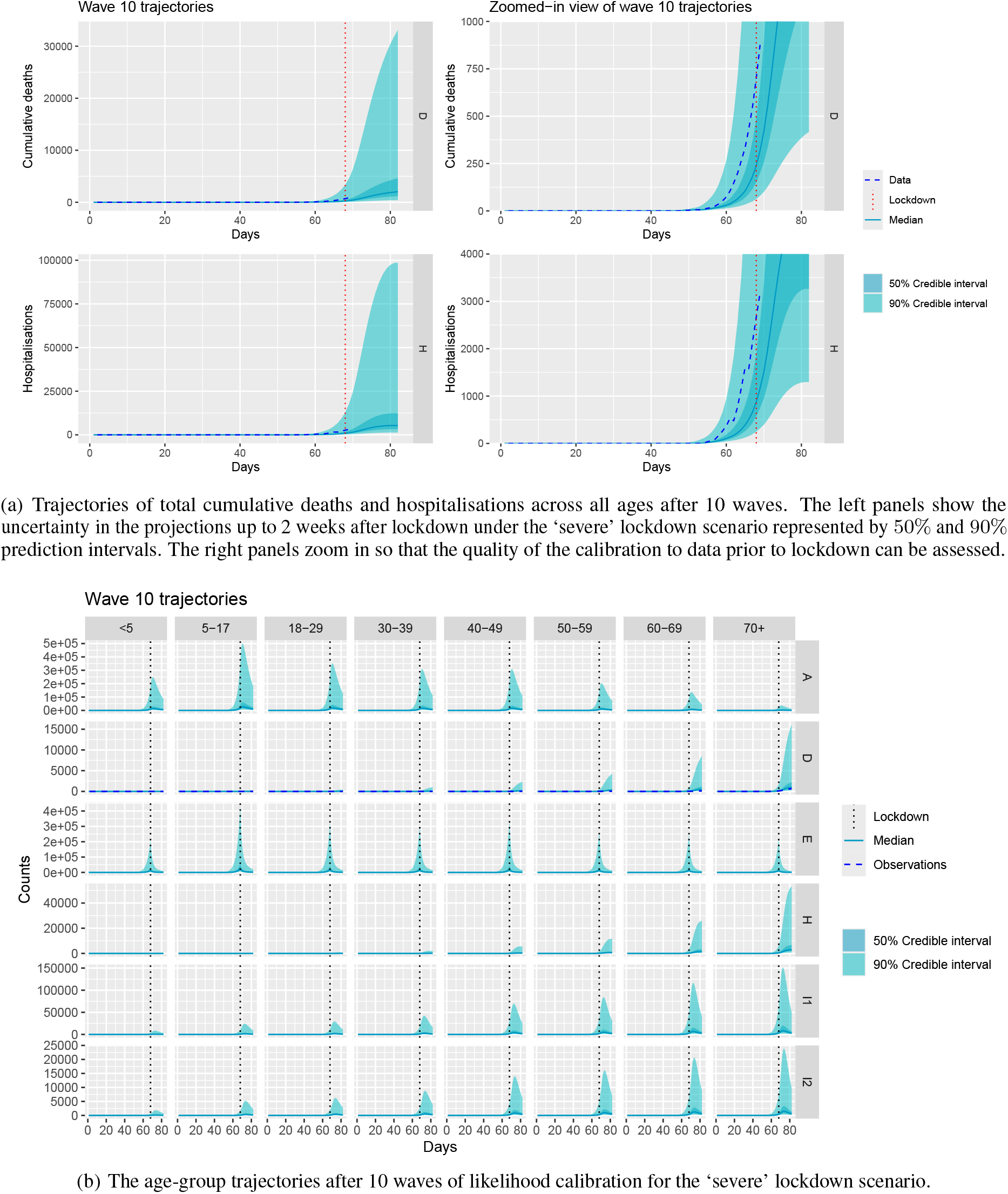
Calibrated and projected COVID-19 epidemic state under the ‘severe’ lockdown scenario.

We see from the plots up to lockdown that, whilst the calibration has worked well, the data trajectory is between the 75^th^ and 95^th^ percentiles of our uncertainty about the true state of the disease. There is nothing amiss with this *per se*, but it is worth noting that our model favours true disease states that are higher than observations given that we expect an under-reporting bias given our choice of Skellam parameters. The observations being above our median trajectory is not evidence of a problem, but it might also be something we see if there was an issue (say with our sampling being biased towards the low end of the potential outbreak). We say more about this in the discussion.

Figure 7 (b) shows the main states of the epidemic. We note that the model predicts high asymptomatic infection rates amongst young people, particularly those of school age. The severity of the disease increases with age and we expect roughly 200,000–300,000 people to have been exposed to the disease from each age-class by lockdown.

### 5.3 Scenario forecasts for lockdown policy

Figure 8 shows probabilistic projections of total deaths from COVID-19 beyond lockdown under the four hypothetical lockdown scenarios. The left plots show the 90% prediction intervals commonly reported in Bayesian analyses (shading) around the median line, and the right plots show the interquartile range. We show both to illustrate that the predictive distributions are heavy tailed (the 95^th^ percentile is an order of magnitude larger than the 75^th^). Note that we can easily do any number of scenarios post-calibration in this way, starting from the calibrated set of particles at lockdown. Figures such as these could help communicate, either to policymakers or to those whom lockdown would affect (the population) the rationale behind any decision making. Figure 9 shows the same projections for hospitalisations. For both deaths and hospitalisations, we note that any lockdown has a significant impact on deaths and on hospitalisations compared to business as usual and that the severity of the lockdown has a smaller impact. This could be down to using the CoMix contact matrix for all three lockdown scenarios. If using our methodology for a new outbreak, further parameters would be introduced to dampen the intergenerational contacts, similar to our *β*_*s*_ and these would be different for each scenario.

**Figure 8:**
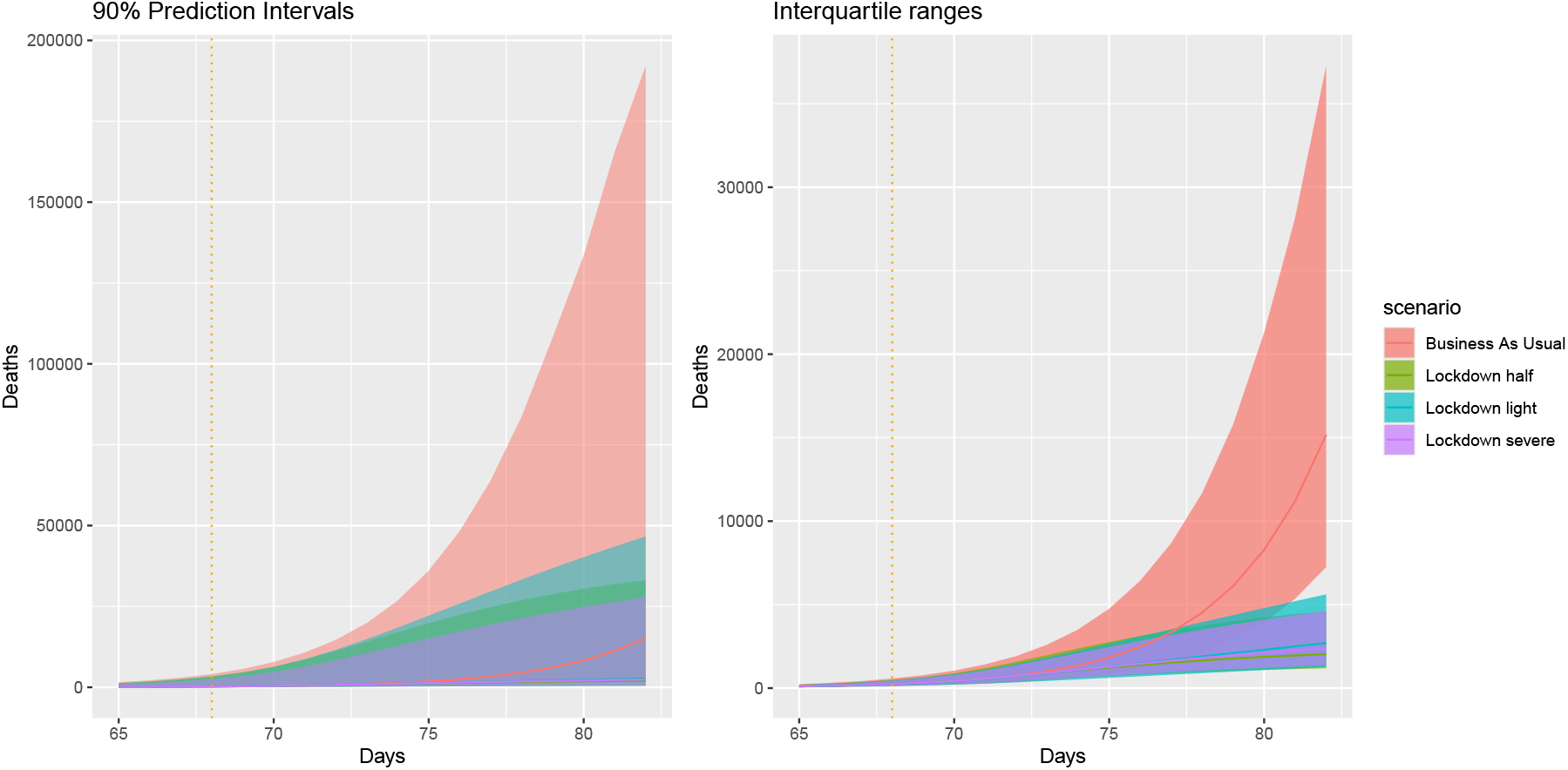
Probabilistic projections of total COVID-19 deaths for 2 weeks after lockdown under the 4 lockdown scenarios explored. The left panel shows the 90% prediction intervals, whilst the interquartile range (or 50% prediction intervals) are shown in the right panel.

**Figure 9:**
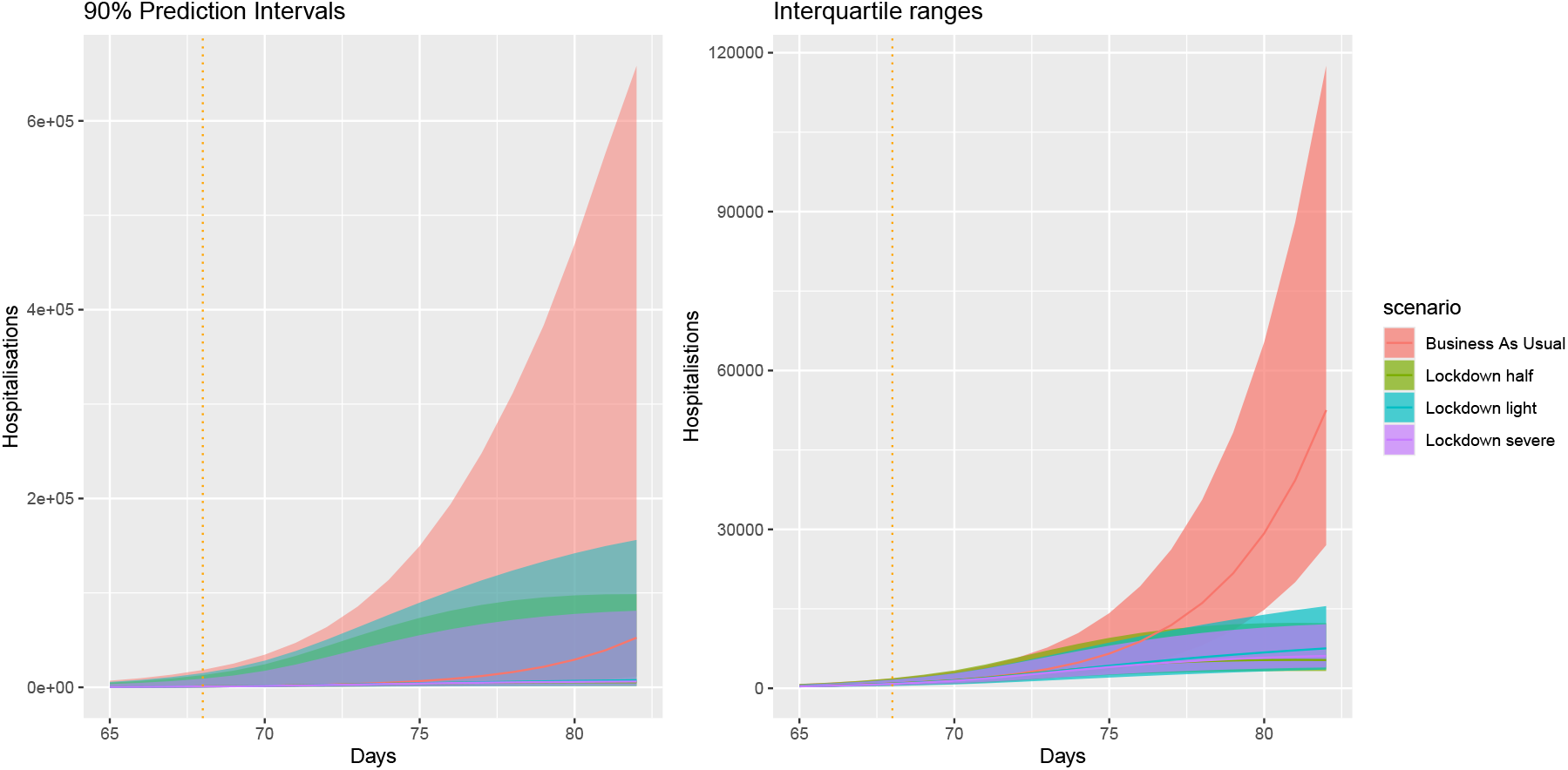
Probabilistic projections of total COVID-19 hospitalisations for 2 weeks after lockdown under the 4 lockdown scenarios explored. The left panel shows the 90% prediction intervals, whilst the interquartile range (or 50% prediction intervals) are shown in the right panel.

To present the type of probabilistic summary we might be able to construct for policymakers considering lockdown (and the severity of the measures to put in place), we consider the probabilities of exceeding an average 1, 000 deaths per day, exceeding the unoccupied bed capacity within the NHS (for England) and exceeding the total NHS-England bed capacity, within the two week period following lockdown for each of our four scenarios. We use Ewbank et al. [2021] for NHS bed capacity, which is given as 141, 000 for 2019/20, with an average bed occupancy of 90.2%. We then use the spare 9.8% of UK beds to represent the unoccupied bed capacity (13, 818). We note that the length of the projections, the types of scenario and the policy-relevant measures are all presented as examples of the things that can be explored with our real-time calibration system, rather than examples of the things that should be (or should have been) considered.

Without some form of restrictions on movement, our forecast for the probability of exceeding an average of 1, 000 deaths per day was 52%, with a 92% chance of breeching spare NHS bed capacity by that time, and a 23% chance that the numbers needing hospital treatment for COVID-19 would exceed the total NHS bed capacity. The lockdown scenarios we explored reduced the probabilities of these extreme events considerably, though even the best-case had a near 2% chance of exceeding NHS capacity. The chances of exceeding the available NHS beds remained around 25% under all lockdown scenarios, suggesting that existing levels of infection were already high enough to require further mitigation of impacts. We note that our probabilities are similar enough for all lockdown scenarios that it is likely that the use of the Co-Mix contact matrix has the dominant effect, rather than our scaling of the forcing of infection.

## 6 Discussion

We have presented a real-time calibration framework for pandemic modelling that is designed to feed into policy support systems (e.g. through the systems used during COVID19 in the UK [Silk et al., 2022, Bowman, 2022]). It is motivated by the knowledge that expensive ID models are changed, sometimes even drastically, from week to week in order to answer policy questions during a pandemic, and that the turnaround time from data being made available to deadlines to feed in to policy support can be as little as one day [Brooks-Pollock et al., 2021]. Our framework embeds structural model discrepancy into all states of the model, with an easy-to-specify parameterisation based on Skellam distributions. The structure and conditional bounds of the joint discrepancy are all automatically derivable from the graphical representation of the model (as presented in Section 3.1). We use particle filtering to estimate the log-likelihood and build our emulation and calibration framework around targeting this expensive object. As the log-likelihood is always 1 dimensional, emulation and multi-wave calibration is readily automatable and the methodology is model-agnostic, meaning that changes made by modellers designed to address questions during the pandemic do not require bespoke-UQ to handle the calibration. Moreover, our use of embedded discrepancy with the data assimilation removes the usual requirement to ‘seed’ the outbreak artificially, and to enable the start of the outbreak to be estimated by the model. It also allows infections to occur externally, perhaps mimicking arrival of infected individuals from other countries.

Our methodology is fundamentally different in spirit to other attempts to use UQ for calibration of ID models, which emulate the ID model outputs and compare them to data. When the ID model is fixed and the turnaround time from model-change to policy support is weeks or months, these methods remain powerful and arguably the go-to (in terms of UQ). However, when the turnaround time is short, the requirement for a new structural discrepancy model and bespoke emulation of potentially novel output fields renders the approach difficult and unreliable for short timescales. Within the ID modelling literature, there are many calibration approaches that quantify many or most uncertainties and even leverage particle filters (e.g. via particle MCMC, Andrieu et al., 2010). These methods can work well, but are computationally intensive, requiring many more runs of the particle filters than our emulator-based approaches, and thus, arguably, making them less scalable to high-resolution spatial models. Current models also do not capture model discrepancy and still require the outbreak to be manually seeded, or at least for the seeding to be estimated along with the other parameters.

We presented an application of our methodology for an age-structured model; however, the UQ problem is of the same complexity for a high-resolution spatial model. There are a number of computational innovations required to deliver the same system over high-resolution spatial ID models, and a number of specific concerns and situations to address. We address those and provide our spatial implementation in Part 2 of this paper, [McKinley et al., 2025], where the same UQ approach, with an automated embedded model-discrepancy and multi-wave likelihood calibration with 1D emulation, is applied and where further challenges surround the data assimilation. Our implementation here used the BPF, but more advanced particle filtering techniques, such as the twisted particle filter [Guarniero et al., 2017], which can look ahead, could be more appropriate, due to the data we assimilate usually being elements at the end of a disease pathway. Future work will explore the adaptation of these kinds of filters to compartmental ID models with embedded model discrepancy. Fast approximate look-ahead filters also show great potential in this area [Whiteley and Rimella, 2021].

There are several further avenues to explore for our own methodology. Our calibration currently returns a uniform sample from the remaining parameter space after a pre-determined number of waves of likelihood calibration. Oakley and Youngman [2017] gave a method for importance sampling from a likelihood emulator to deliver a posterior distribution over the parameters. The particle filter made this approach infeasible for us, as the method requires an emulator with almost no variance to work well, and our filtering only estimates the log-likelihood and still has large noise even with 500 particles. However, interest remains in estimating a posterior over the parameters after some number of waves. We might also consider Bayesian optimisation with our emulators to find the MLE, making the estimation of the NROY set via (11) robust in later waves once the optimisation had converged. Our application used 10 waves for each calibration, but a stopping rule might be derived. One stopping rule could involve testing for convergence of the percentage of space ruled out, another might consider when the emulator variance is smaller than the variance of the particle filter. These remain ideas for further work.

Gaussian process methods, such as ours, often do not work well with large numbers (say *>* 10) of parameters. History matching type approaches have typically tackled this by using subsets of the parameters deemed most ‘active’ [Vernon et al., 2010]; however, these approaches still struggle if many parameters are active. These considerations motivated our choice of log-linear parameterisation for age-related progression probabilities. However, recent advances in estimating active subspaces [Constantine et al., 2014, Kellin Rumsey and Wiel, 2024] may open the possibility to calibrate even large numbers of parameters automatically, allowing every age-class to have its own transition parameters. This is the subject of forthcoming work.

Our particular parameterisations of the Skellam discrepancy and observation distributions represented a modelling choice we made to support our narrative, with the values we gave for the parameters justified based on expert judgement. However, both the form and the choice of parameters can be given further attention. We mentioned exploring non-linear (in model state) parameterisations, which can certainly be done. We can also make the parameters of the Skellam distributions themselves calibratable, if the modeller would prefer not to set them. This would only be a reasonable thing to do if enough prior information were included to overcome identifiability problems [Brynjarsdóttir and O’Hagan, 2014]. The heuristics we used to fix the parameters could easily be re-purposed to deliver strong prior information for identifying the discrepancy parameters via the calibration. Our data error modelling is relatively unstructured and delivers a simple parameterisation for under-reporting. We stress here that our method is trivially applied to complex data models, such as those developed by epidemiologists and modellers during the pandemic to attempt to capture the biases and uncertainties in the data from different streams. The data model is used to weight particles during the filtering, and hence our method is somewhat independent from any specific data modelling choices.

Delivering an operational real-time calibration system for high-resolution infectious disease models is not something that can be trivially done during the outbreak of the next pandemic. The system itself needs to be delivered before the next pandemic arrives, though this does not require a specific model. A modelling framework like, for example, the one provided by the open source framework Metawards [Woods et al., 2022], would need to be enabled with an advanced ‘data assimilation’ mode, capable of introducing model discrepancy and returning the log-likelihood for different parameter values and for a large enough number of particles to deliver accurate estimates. The emulation/calibration framework can sit alongside this and be relatively lightweight, but HPC will likely be needed to perform advanced particle filtering for a high-resolution spatial model. We have taken considerable steps towards such a framework in Part 2 of this paper, but there is more work to be done. Without that work, or considerable innovation with another approach, we may face our next pandemic once again without the advantages that tools from Uncertainty Quantification can provide for speeding up the estimation of high-resolution ID models.

## Data Availability

All data produced by the present study are available upon reasonable request to the authors

