## Supplementary material for "On real-time calibrated prediction for complex model-based decision support in pandemics: Part 1"

### S1 Truncated discrete Gaussian distributions

The probability mass function for an arbitrary $X \sim tN^D\left(\mu, \sigma^2, \mathrm{LB}, \mathrm{UB}\right)$ is given by:

$$P\left(X = x\right) = \frac{\int_{x-0.5}^{x+0.5} \frac{1}{\sigma\sqrt{2\pi}} \exp\left(-\frac{1}{2}\left[\frac{x^*-\mu}{\sigma}\right]^2\right) dx^*}{\int_{\mathrm{LB}-0.5}^{\mathrm{UB}+0.5} \frac{1}{\sigma\sqrt{2\pi}} \exp\left(-\frac{1}{2}\left[\frac{w^*-\mu}{\sigma}\right]^2\right) dw^*} \quad x = \mathrm{LB}, \ldots, \mathrm{UB}; \sigma^2 > 0, \tag{S.1}$$

which is straightforward to evaluate using standard libraries. To sample from (S.1) we can use the method developed by Chopin (2011) to efficiently sample a continuous latent variable $Z^* \sim tN\left(0, 1, \frac{\mathrm{LB}-\mu-0.5}{\sigma}, \frac{\mathrm{UB}-\mu+0.5}{\sigma}\right)$ before setting $X = \lfloor \sigma Z^* + \mu + 0.5 \rfloor$. For full details please see Section S1.1.

#### S1.1 Sampling from a discrete truncated Gaussian

Let $Z^* \sim tN\left(0, 1, a, b\right)$ be a truncated standard Gaussian random variable, truncated in the region $(a, b)$ for some $-\infty < a < b < \infty$, with probability density function

$$f_{Z^*}\left(z^*\right) = \frac{\frac{1}{\sqrt{2\pi}} \exp\left(-\frac{(z^*)^2}{2}\right)}{\int_a^b \frac{1}{\sqrt{2\pi}} \exp\left(-\frac{(u^*)^2}{2}\right) du^*} \quad a < z^* < b. \tag{S.2}$$

The method of Chopin (2011) provides a fast and efficient way to produce random samples from this distribution. Hence, if we take a random sample, $Z^* \sim tN\left(0, 1, \frac{\mathrm{LB}-\mu-0.5}{\sigma}, \frac{\mathrm{UB}-\mu+0.5}{\sigma}\right)$, where $\mathrm{LB} < \mathrm{UB}$ and $\mathrm{LB}, \mathrm{UB} \in \mathbb{Z}$, and then set $X = \lfloor \sigma Z^* + \mu + 0.5 \rfloor$, then $X$ is a random sample from a truncated discrete Gaussian distribution with probability mass function:

$$P\left(X = x\right) = \frac{\int_{x-0.5}^{x+0.5} \frac{1}{\sigma\sqrt{2\pi}} \exp\left(-\frac{1}{2}\left[\frac{x^*-\mu}{\sigma}\right]^2\right) dx^*}{\int_{\mathrm{LB}-0.5}^{\mathrm{UB}+0.5} \frac{1}{\sigma\sqrt{2\pi}} \exp\left(-\frac{1}{2}\left[\frac{w^*-\mu}{\sigma}\right]^2\right) dw^*} \quad x = \mathrm{LB}, \ldots, \mathrm{UB}; \sigma^2 > 0; -\infty < \mu < \infty, \tag{S.3}$$

and hence $X \sim tN^D\left(\mu, \sigma, \mathrm{LB}, \mathrm{UB}\right)$ as required.

To prove this, consider a random variable $X^* = \sigma Z^* + \mu$ where $Z^* \sim tN\left(0, 1, \frac{\mathrm{LB}-\mu-0.5}{\sigma}, \frac{\mathrm{UB}-\mu+0.5}{\sigma}\right)$. Since $\sigma > 0$ we can use the general monotonic transformation equation to derive the p.d.f. of $X^*$; hence:

$$\begin{aligned} f_{X^*}\left(x^*\right) &= f_{Z^*}\left(\frac{x^*-\mu}{\sigma}\right)\left|\frac{d}{dx^*}\left(\frac{x^*-\mu}{\sigma}\right)\right| \\ &= \frac{\frac{1}{\sqrt{2\pi}} \exp\left(-\frac{\left(\frac{x^*-\mu}{\sigma}\right)^2}{2}\right)}{\int_{\frac{\mathrm{LB}-\mu-0.5}{\sigma}}^{\frac{\mathrm{UB}-\mu+0.5}{\sigma}} \frac{1}{\sqrt{2\pi}} \exp\left(-\frac{(u^*)^2}{2}\right) du^*} \times \frac{1}{\sigma} \qquad \text{from (S.2)} \end{aligned} \tag{S.4}$$

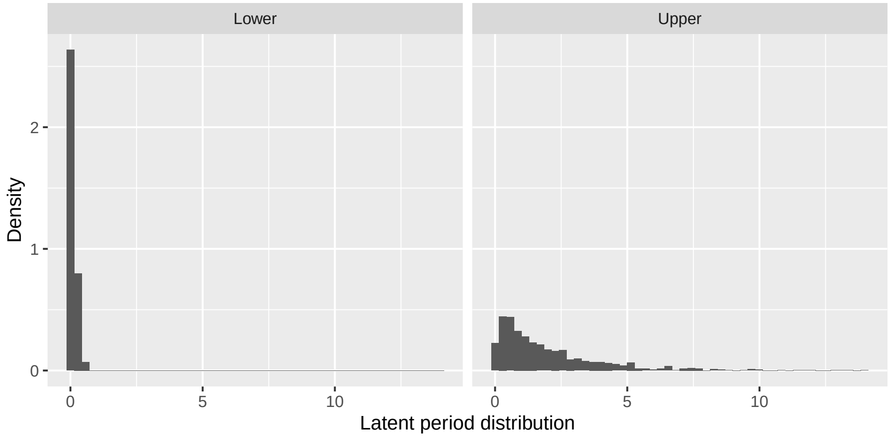


Figure S1: The latent period distributions at the lower and upper bounds of the parameter ranges for $T_E$

Letting $w^* = \sigma u^* + \mu$ (and hence $dw^* = \sigma du^*$), and then performing a change-of-variables in the integral in (S.4) gives:

$$f_{X^*}(x^*) = \frac{\frac{1}{\sigma\sqrt{2\pi}} \exp\left(-\frac{(x^*-\mu)^2}{2\sigma^2}\right)}{\int_{\text{LB}-0.5}^{\text{UB}+0.5} \frac{1}{\sigma\sqrt{2\pi}} \exp\left(-\frac{1}{2}\left[\frac{w^*-\mu}{\sigma}\right]^2\right) dw^*} \qquad \text{LB} - 0.5 < x^* < \text{UB} + 0.5; \sigma^2 > 0; -\infty < \mu < \infty. \quad \text{(S.5)}$$

Therefore $X^* \sim N\left(\mu, \sigma^2, \text{LB} - 0.5, \text{UB} + 0.5\right)$, and hence if $X = \lfloor X^* + 0.5 \rfloor$ then $P(X = x) = P(x - 0.5 < X^* < x + 0.5)$ for $x = \text{LB}, \ldots, \text{UB}$ and as such $X$ has probability mass function (S.3) as required.

### S1.2 Prior elicitation

We have derived plausible parameter bounds from data and the literature. When the original elictation was done we used data from the original 2020 pre-prints of the referenced texts below, but include the published versions in the descriptions below.

$$\begin{aligned}
R_0 &: (2, 4.5) \qquad \text{from co-authors Danon and Challen}\\
\nu_A &: (0, 1)\\
T_E &: (0.1, 2)\\
T_P &: (1.2, 3)\\
T_{I_1} &: (2.8, 4.5)\\
T_{I_2} &: (0.0001, 0.5)\\
\alpha_{EP}, \alpha_{I_1H}, \alpha_{I_1D}, \alpha_{HD}, \eta &: \text{drawn from distribution as described below}\\
\eta_I &: (0.5, 2)\\
\eta_H &: (0.5, 2)\\
\alpha_{T_H}, \eta_{T_H} &: \text{drawn from distribution as described below}\\
\beta_s &: (0, 1)
\end{aligned}$$

For the mean latent period, $T_E$, we set the upper bound such that the latent period distribution has an upper tail probability that is consistent with the *generation interval* distribution from Challen et al. (2022) (Figure 4 in that paper). This provides a conservative upper bound, since the generation time is equivalent to $T_E + T_P$ in our model. We choose a lower bound such that it is possible to have a short, but non-zero latent period. The latent period distributions at the lower and upper bounds of the parameter ranges for $T_E$ are shown in Figure S1.

To generate ranges for the length of the pre-symptomatic period, $T_P$, we note that the *incubation period* is equivalent to $T_E + T_P$ in our model, and hence through simulation we generated incubation period distributions

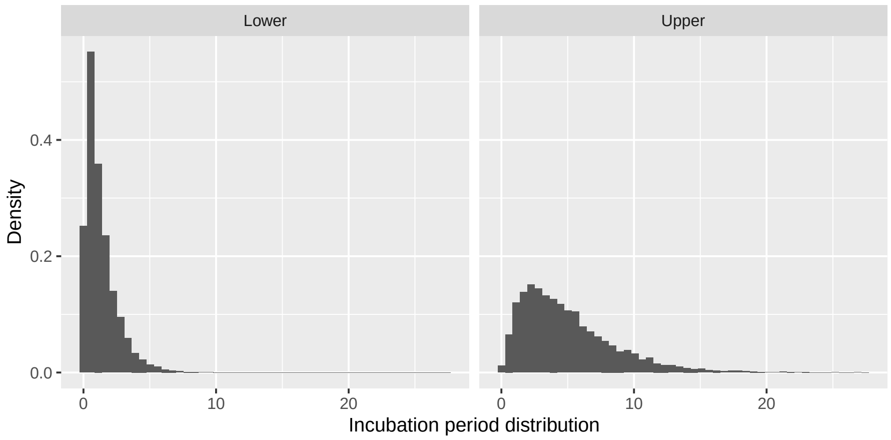


Figure S2: The incubation period distributions at the lower and upper bounds of the parameter ranges for $T_E$ and $T_P$

at the lower and upper bounds of $T_E$, and chose bounds for $T_P$ such that at the lower bounds of $T_E$ and $T_P$ the incubation period distribution looks like the lower-mean incubation period distribution in Challen et al. (2022) (Figure 3A in that paper), and at the upper bounds of $T_E$ and $T_P$ the incubation period distribution looks like the higher-mean incubation period distribution in Challen et al. (2022) (Figure 3B in that paper). Figure S2 shows the incubation period distributions at the lower and upper bounds of the parameter ranges for $T_E$ and $T_P$.

To generate ranges for the length of the initial infectious period, $T_{I_1}$, we note that this is equivalent to the *time-from-onset-to-admission* in our model, and hence through simulation chose bounds for $T_{I_1}$ such that its distribution was approximately consistent with the time-from-onset-to-admission in Challen et al. (2022) (Figure 7A in that paper), and also that the time-from-infection-to-admission, $T_E + T_P + T_{I_1}$, was consistent with the time-from-infection-to-admission in Challen et al. (2022) (Figure 7B in that paper), at both the lower and upper bounds for $T_E$ and $T_P$. Figure S3A shows the *time-from-onset-to-admission* ($T_{I_1}$) distribution at the lower and upper bounds of $T_E$ and $T_P$, and Figure S3B shows the derived *time-from-infection-to-admission* distribution at the lower and upper bounds of $T_E$, $T_P$ and $T_{I_1}$.

For the post-onset infectious period distribution, $T_{I_2}$, we noted that Cevik et al. (2021) found no live virus beyond day 9 of symptom onset. As such, we used simulation to find upper and lower bounds for $T_{I_2}$. The upper bound of $T_{I_2}$ was chosen so that when $T_{I_1}$ was at its lower bound, the probability that the infectivity time post onset ($T_{I_1} + T_{I_2}$) was >9 days was $\approx 0.05$. Conversely, when $T_{I_1}$ was at its upper bound, then even for very low values of $T_{I_2}$ we were unable to get upper tail probabilities for $T_{I_1} + T_{I_2}$ of less than $\approx 0.13$, hence we set a lower bound for $T_{I_2}$ of 0.0001. At the upper bound for $T_{I_1}$ and $T_{I_2}$ these tail probabilities were $\approx 0.15$. Figure S4 shows the post-onset infectious period distribution ($T_{I_2}$) at the lower and upper bounds of $T_{I_1}$ and $T_{I_2}$.

For the hospital stay times we used data from the CHESS study NHS Digital (2021). The data were supplied after anonymisation under strict data protection protocols agreed between the University of Exeter and Public Health England. The ethics of the use of these data for these purposes was agreed by Public Health England with the UK government SPI-M(O)/SAGE committees. The data is now available from various trusted research environments under the new name or SARI-Watch, including via NHS England Data Access Environment, the UK Longitudinal Linkage Collaboration, or the Clinical Practice Research Datalink. Figure S5 shows the distribution of stay times by age-class, along with the best fitting exponential model in each case (estimated using maximum likelihood).

If we plot the log(mean hospital stay time) from the fitted exponential models against the midpoints of each age-category, then we get an approximate increasing linear relationship with age (Figure S6). To capture estimation uncertainties more robustly, we built a hierarchical Bayesian model (fitted using MCMC implemented in the NIMBLE package in R—de Valpine et al., 2017) to estimate both the mean hospital stay times in each age category, and then simultaneously the slope and intercept from Gaussian linear regression model fitted to the log(mean hospital stay time) from the exponential models. Figure S6 shows the log(mean hospital stay time) estimates from the non-Bayesian models (fitted independently to each age-distribution) against the fitted line from the hierarchical Bayesian model, with 99% credible intervals.

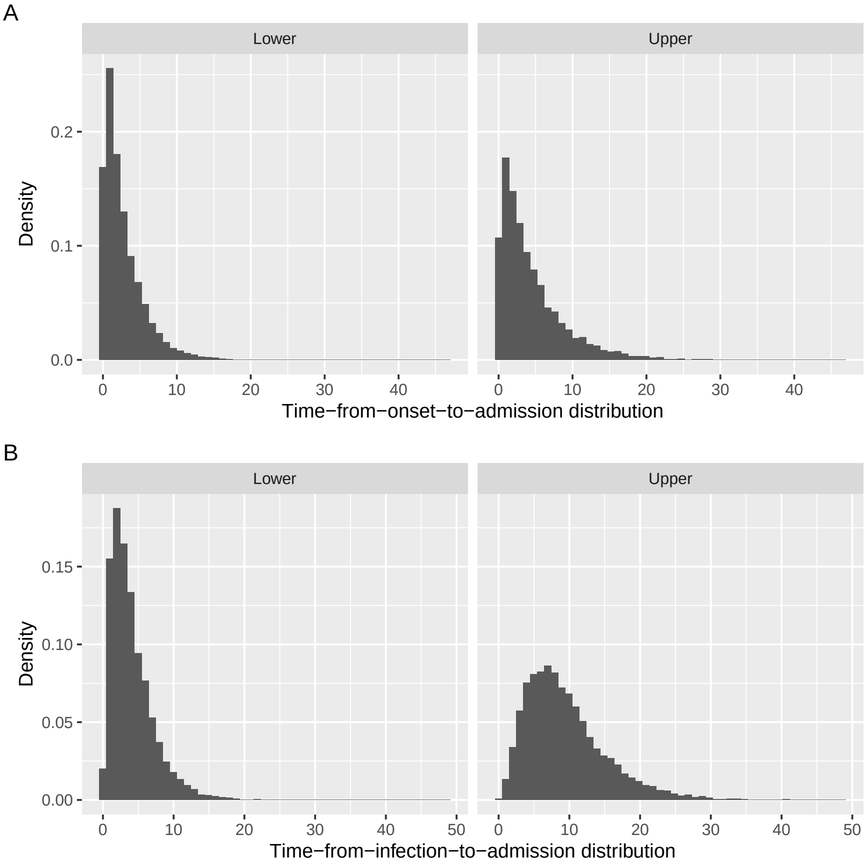


Figure S3: A: the *time-from-onset-to-admission* ($T_{I_1}$) distribution at the lower and upper bounds of $T_E$ and $T_P$; B: the *time-from-infection-to-admission* distribution at the lower and upper bounds of $T_E$, $T_P$ and $T_{I_1}$

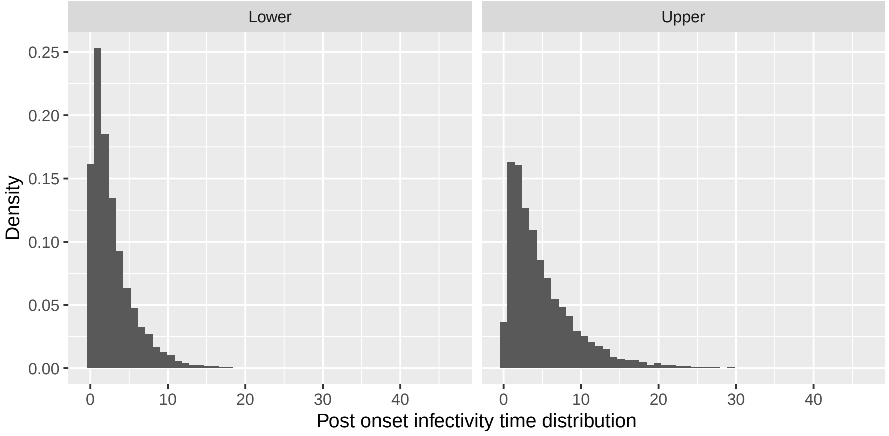


Figure S4: Post-onset infectious period distribution ($T_{I_2}$) at the lower and upper bounds of $T_{I_1}$ and $T_{I_2}$

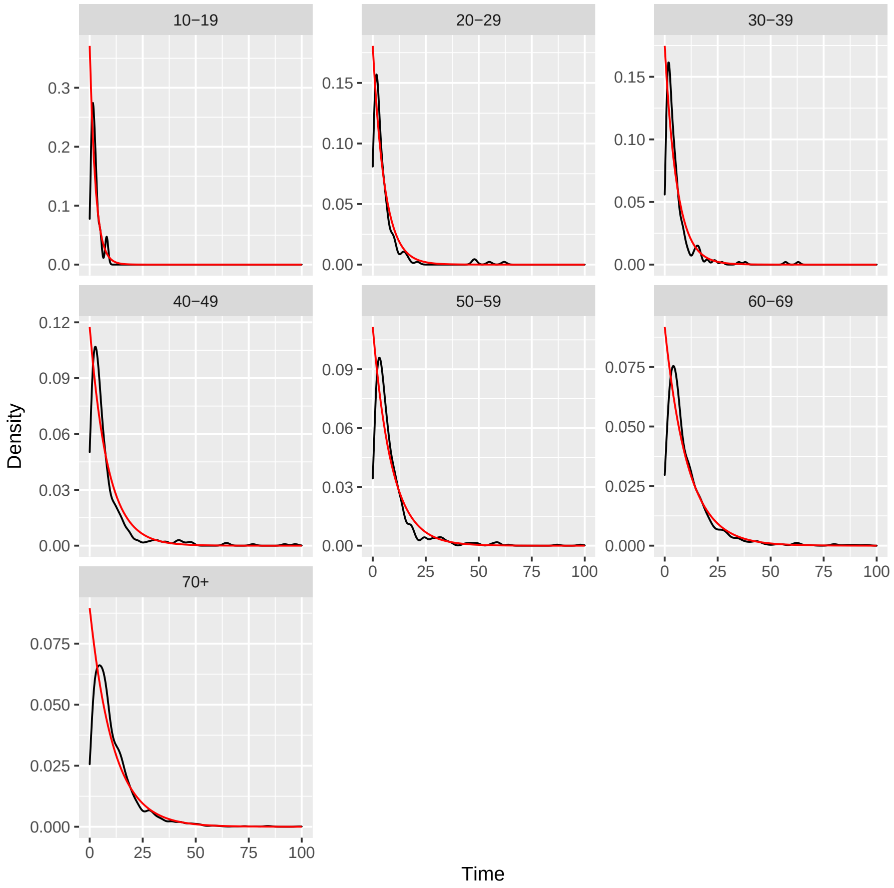


Figure S5: Hospital stay times by age-class from the CHESS study. The kernel density estimates of the observed distributions are shown by the black lines, and the red lines correspond to the best-fitting exponential model in each case

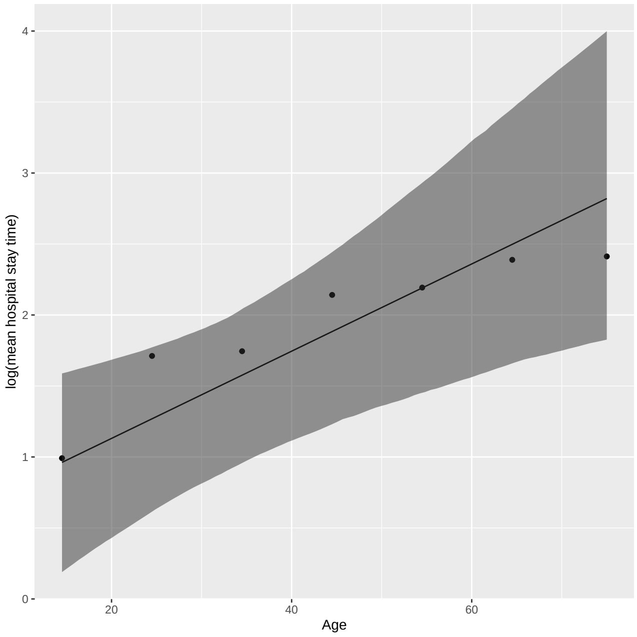


Figure S6: log(mean hospital stay time) estimates from the non-Bayesian models (black points) against the fitted line from the hierarchical Bayesian model (black line), with 99% credible intervals shown by the grey ribbon

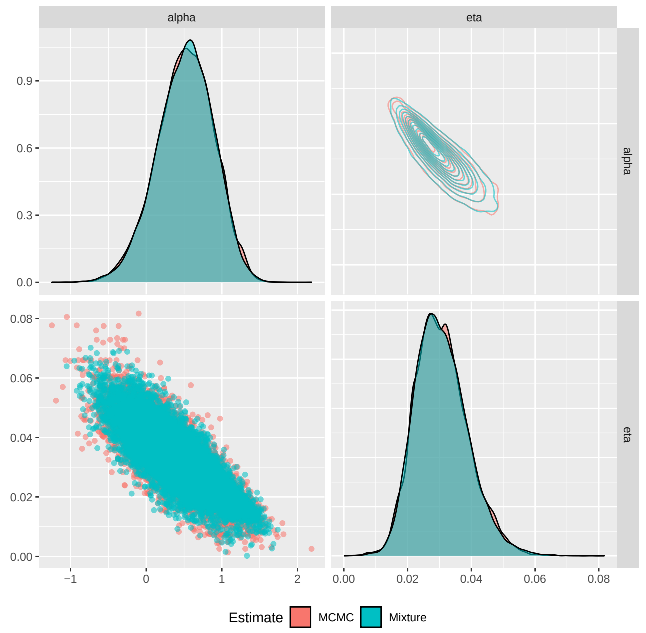


Figure S7: Kernel density estimate for the posterior distributions for $\alpha$ and $\eta$ from the model fitted in Figure S6 against the kernel density estimate from an approximating finite Gaussian mixture model

The intercept and slope of this line provide the parameters we use to determine the age-structured hospital stay times. To generate the initial design for the history matching, we draw from the joint posterior distribution using a space-filling design for these parameters. We do this by generating large numbers of samples from the posterior, and then sub-sample these using a *maximin* design to get the appropriate coverage. To avoid having to run the MCMC each time we want more design points, we approximate the posterior distribution using a truncated finite Gaussian mixture model (FMM), fitted using the `mclust` package in R Scrucca et al. (2023). The posterior distributions from the MCMC and the approximating FMM are shown in Figure S7.

For the probabilities of transitioning along different pathways by age, we used two sources of information to generate plausible ranges to use as initial inputs into the calibration. We use model estimates of the *infection fatality risk* and the *probability of hospitalisation given infection* from Verity et al. (2020), and also some data from the Centers for Disease Control (CDC).

The estimates of the infection fatality risk and the probability of hospitalisation given infection from the CDC are obtained in the following way.

1. The cumulative number of cases and deaths for each age group by 26/02/2021 were downloaded from https://covid.cdc.gov/covid-data-tracker/#demographics. 2. We use the cumulative hospitalisation proportion (per 100,000 population) on 20/02/2021 (available from https://gis.cdc.gov/grasp/COVIDNet/COVID19_3.html), and the US population by age (available from https://www.statista.com/statistics/241488/population-of-the-us-by-sex-and-age/) to estimate the number of hospitalisations for the entire US population. 3. We obtain the number of in hospital deaths by age group from https://data.cdc.gov/dataset/NVSS-Provisional-COVID-19-Deaths-by-Place-of-Death/4va6-ph5s/data, by filtering the data for the entire US using the category ”Place of death” / ”Healthcare setting, inpatient”.

If we plot the log(probability of hospitalisation given infection), and the log(infection fatality risk) against the midpoints of each age-category, then we get an approximate increasing linear relationship with age in all cases, and after discussions we have decided to use log-linear relationships between the probabilities of transitions along certain pathways, and age, with the same slope parameter in each case, constrained such that there is always an increasing risk of more severe outcomes with age. We allow the intercepts to vary between the different transition pathways. Hence we have five parameters to calibrate: the intercepts $\alpha_{EP}$, $\alpha_{I_1D}$, $\alpha_{I_1H}$, $\alpha_{HD}$ and the common slope $\eta$.

To derive plausible ranges for these parameters, we used an ad-hoc MCMC approach based on simulated data sets. Firstly, we took the infection fatality risks and probabilities of hospitalisation estimates from Verity et al. (2020), and for each age-class we generated a pseudo-data set by randomly sampling from a binomial distribution

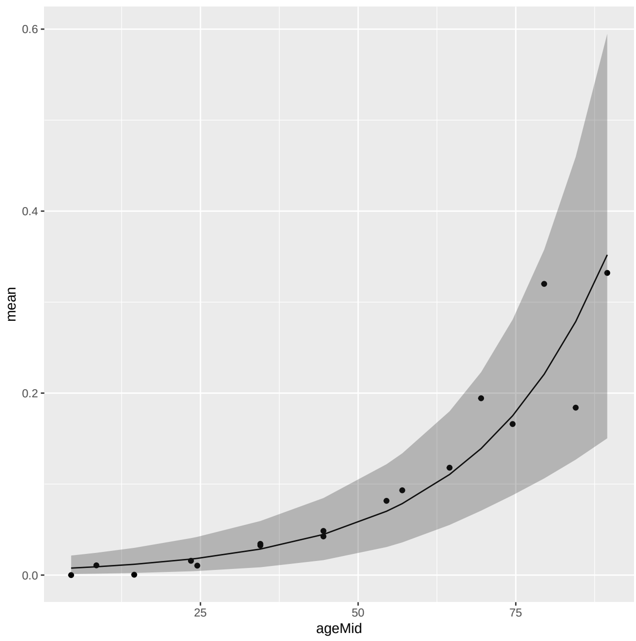


Figure S8: Probability of hospitalisation given infection estimates from both the Verity et al. (2020) and CDC data (black points) against the posterior mean (black line) and 99% prediction intervals (grey ribbon) generated from the union of posterior samples

with 100 trials and a probability of success given by the relevant estimate. This gave us age-specific counts for hospitalisations ($y_{iH}$) and death *given* infection ($y_{iD}$) given $n_i = 100$ "trials" for age-classes $i$. We then fitted a model to this pseudo-data set, targeting a posterior of the form:

$$\pi\left(\alpha_{EP}, \alpha_{I_1D}, \alpha_{I_1H}, \alpha_{HD}, \eta \mid \mathbf{y}, \mathbf{n}\right) \propto \tag{S.6}$$
$$\pi\left(\mathbf{y} \mid \alpha_{EP}, \alpha_{I_1D}, \alpha_{I_1H}, \alpha_{HD}, \eta, \mathbf{n}\right) \pi\left(\alpha_{EP}, \alpha_{I_1D}, \alpha_{I_1H}, \alpha_{HD}, \eta\right). \tag{S.7}$$

The likelihood component, $\pi\left(\mathbf{y} \mid \alpha_{EP}, \alpha_{I_1D}, \alpha_{I_1H}, \alpha_{HD}, \eta, \mathbf{n}\right)$, depends on a set of latent variables, $\mathbf{x}$, and can be written as:

$$\pi\left(\mathbf{y} \mid \alpha_{EP}, \alpha_{I_1D}, \alpha_{I_1H}, \alpha_{HD}, \eta, \mathbf{n}\right) = \int_{\mathcal{X}} \pi\left(\mathbf{y} \mid \mathbf{x}, \mathbf{n}\right) \pi\left(\mathbf{x} \mid \alpha_{EP}, \alpha_{I_1D}, \alpha_{I_1H}, \alpha_{HD}, \eta\right) d\mathcal{X},$$

where $\mathcal{X}$ is the (multidimensional) parameter space for the latent states. Here the latent states correspond to the individuals transitioning down the different pathways in the model, and $\pi\left(\mathbf{y} \mid \mathbf{x}, \mathbf{n}\right)$ is a binomial probability mass function based on probabilities derived from $\mathbf{x}$ (here we assume that given $\mathbf{x}$, the data $y_{iH}$ and $y_{iD}$ are independent). To approximate the integral we used Monte Carlo simulation such that

$$\hat{\pi}\left(\mathbf{y} \mid \alpha_{EP}, \alpha_{I_1D}, \alpha_{I_1H}, \alpha_{HD}, \eta, \mathbf{n}\right) = \frac{1}{M} \sum_{j=1}^{M} \pi\left(\mathbf{y} \mid \mathbf{x}_j, \mathbf{n}\right),$$

where (with a slight abuse of notation) $x_j \sim \pi\left(\mathbf{x} \mid \alpha_{EP}, \alpha_{I_1D}, \alpha_{I_1H}, \alpha_{HD}, \eta\right)$ gives the number of individuals from 100,000 who end up running through each pathway, and for age-class $i$ and count $y_i$: $\pi\left(y_i \mid x_i, n\right) \sim \text{Bin}\left(100, x_i/100,000\right)$ with $x_i$ the relevant simulated count. We used this estimator in place of the true likelihood in a Metropolis-Hastings algorithm to get an approximate "posterior". We found $M = 1$ simulation was sufficient, but we had to run long chains to get good mixing.

We repeated this approach for 10 pseudo-data sets derived from the Verity et al. (2020) estimates, and also 10 pseudo-data sets derived from the CDC data. The *union* of these posterior samples across all 20 data sets provided our plausible ranges.

Figure S8 shows the probability of hospitalisation given infection estimates from both the Verity et al. (2020) and CDC data against the 99% prediction intervals generated from our union of posterior samples.

Figure S9 shows the probability of death given infection estimates from both the Verity et al. (2020) and CDC data against the 99% prediction intervals generated from our union of posterior samples.

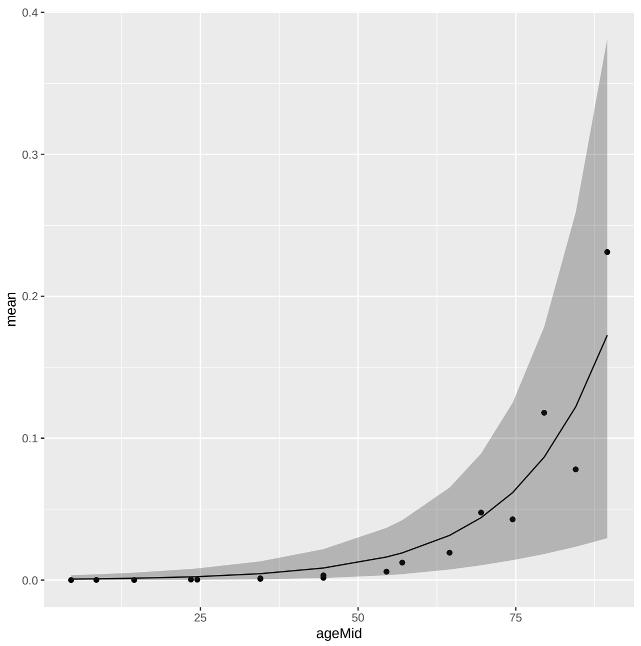


Figure S9: Probability of death given infection estimates from both the Verity et al. (2020) and CDC data (black points) against the posterior mean (black line) and 99% prediction intervals (grey ribbon) generated from the union of posterior samples

Similarly to the approach for the hospital stay parameters, for our input designs we draw from the union of posterior distributions using a space-filling sampling design generated from a large set of posterior samples. To avoid having to run the MCMC each time, we approximate the posterior distribution using a truncated finite Gaussian mixture model, fitted using the `mclust` package in R Scrucca et al. (2023). The posterior distributions from the MCMC and the approximating FMM are shown in Figure S10. In practice we also require checks that $\eta > 0$ and that for any combination of parameters the probabilities of transition in all age-classes are $< 1$, and also that any multinomial probabilities sum to one (so we reject any proposal where $p_{I_1H} + p_{I_1D} > 1$ for example).

R (R Core Team, 2022) packages used in this work include: `mclust` (Scrucca et al., 2023), `tidyverse` (Wickham et al., 2019), `GGally` (Schloerke et al., 2024), `MASS` (Venables and Ripley, 2002), `hmer` (Iskauskas et al., 2024), `fields` (Nychka et al., 2021), `dgpsi` (Ming and Williamson, 2024), `patchwork` (Pedersen, 2022), `Rcpp` (Eddelbuettel and François, 2011), `RcppArmadillo` (Eddelbuettel and Sanderson, 2014) and `lhs` (Carnell, 2024) and `NIMBLE` (de Valpine et al., 2017). All simulator model runs were performed on a MacBook Pro with an M1 Pro chip and 16GB of RAM. The code for replicating the experiments in Part 1 is available on GitHub: https://github.com/XiaoyuHy/uq4covid.github.io.

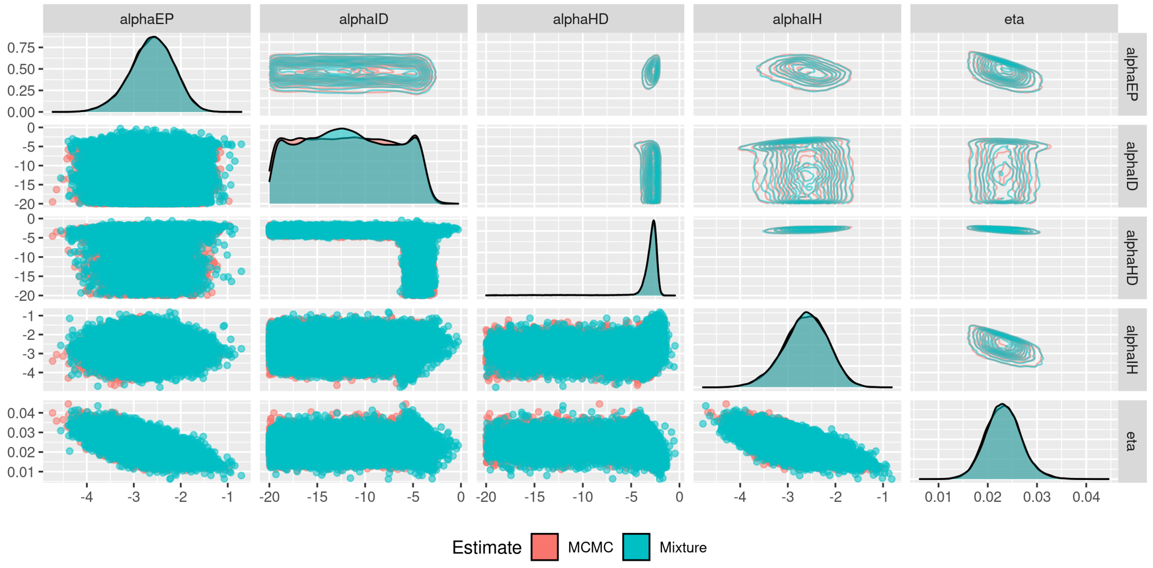


Figure S10: Kernel density estimate for the posterior distributions for $\alpha_{EP}$, $\alpha_{I_1 D}$, $\alpha_{I_1 H}$, $\alpha_{HD}$, $\eta$ from the union of posterior distributions, against the kernel density estimate from an approximating finite Gaussian mixture model

Eddelbuettel, D. and François, R. (2011), "Rcpp: Seamless R and C++ Integration," *Journal of Statistical Software*, 40, 1–18.

Eddelbuettel, D. and Sanderson, C. (2014), "RcppArmadillo: Accelerating R with high-performance C++ linear algebra," *Computational Statistics and Data Analysis*, 71, 1054–1063.

Iskauskas, A., Vernon, I., Goldstein, M., Scarponi, D., McCreesh, N., McKinley, T. J., and White, R. G. (2024), "Emulation and history matching using the `hmer` package," *Journal of Statistical Software*, 109, 1–48.

Ming, D. and Williamson, D. (2024), *dgpsi: An R package powered by Python for modelling linked deep Gaussian processes*, r package version 2.4.0.

NHS Digital (2021), "SGSS and CHESS data (https://digital.nhs.uk/about-nhs-digital/corporate-information-and-documents/directions-and-data-provision-notices/data-provision-notices-dpns/sgss-and-sari-watch-data (accessed Apr 29, 2025))," .

Nychka, D., Furrer, R., Paige, J., and Sain, S. (2021), "fields: Tools for spatial data," R package version 16.3.

Pedersen, T. L. (2022), *patchwork: The Composer of Plots*, R package version 1.1.2.

R Core Team (2022), *R: A Language and Environment for Statistical Computing*, R Foundation for Statistical Computing, Vienna, Austria.

Schloerke, B., Cook, D., Larmarange, J., Briatte, F., Marbach, M., Thoen, E., Elberg, A., and Crowley, J. (2024), *GGally: Extension to 'ggplot2'*, R package version 2.2.1, https://github.com/ggobi/ggally.

Scrucca, L., Fraley, C., Murphy, T. B., and Raftery, A. E. (2023), *Model-Based Clustering, Classification, and Density Estimation Using mclust in R*, Chapman and Hall/CRC.

Venables, W. N. and Ripley, B. D. (2002), *Modern Applied Statistics with S*, New York: Springer, 4th ed., iSBN 0-387-95457-0.

Verity, R., Okell, L. C., Dorigatti, I., Winskill, P., Whittaker, C., Imai, N., Cuomo-Dannenburg, G., Thompson, H., Walker, P. G. T., Fu, H., Dighe, A., Griffin, J. T., Baguelin, M., Bhatia, S., Boonyasiri, A., Cori, A., Cucunubá, Z., FitzJohn, R., Gaythorpe, K., Green, W., Hamlet, A., Hinsley, W., Laydon, D., Nedjati-Gilani, G., Riley, S., van Elsland, S., Volz, E., Wang, H., Wang, Y., Xi, X., Donnelly, C. A., Ghani, A. C., and Ferguson, N. M. (2020), "Estimates of the severity of coronavirus disease 2019: a model-based analysis," *Lancet Infectious Diseases*, 20, 669–677.

Wickham, H., Averick, M., Bryan, J., Chang, W., McGowan, L. D., François, R., Grolemund, G., Hayes, A., Henry, L., Hester, J., Kuhn, M., Pedersen, T. L., Miller, E., Bache, S. M., Müller, K., Ooms, J., Robinson, D., Seidel, D. P., Spinu, V., Takahashi, K., Vaughan, D., Wilke, C., Woo, K., and Yutani, H. (2019), "Welcome to the tidyverse," *Journal of Open Source Software*, 4, 1686.
